# Pharmacological enhancement of glymphatic function in humans increases the clearance of Alzheimer’s disease-related proteins

**DOI:** 10.64898/2026.03.10.26348048

**Authors:** Paul Dagum, Brieann C. Satterfield, Laurent Giovangrandi, T. Robert Feng, Alejandro Corbellini, Kevin Yarasheski, Brendan P. Lucey, Hans Van Dongen, Jeffrey J. Iliff, Albert T. Cheung

## Abstract

Alzheimer’s disease (AD) is characterized by the mis-aggregation of amyloid β (Aβ) and tau, which is proposed to be driven by impaired Aβ and tau clearance. While sleep-active glymphatic transport contributes to the clearance of Aβ and tau in humans, studies have yet to demonstrate that it is possible to enhance glymphatic transport in humans and that augmenting glymphatic transport improves the clearance of Aβ and tau from the human brain. In two cross-over clinical trials in healthy older adults, we demonstrated that a fixed-dose combination therapy of intravenous dexmedetomidine (0.7 μg/kg/h) and 10 mg oral midodrine (ACX-02), that suppressed central noradrenergic tone while maintaining systemic arterial pressure, increased EEG slow waves, enhanced cerebrovascular pulsatility, and reduced parenchymal resistance to perivascular fluid flow, that have shown to be key determinants of glymphatic transport. Dynamic shifts in plasma mass balance indices of clearance within the brain demonstrated that pharmacological enhancement of glymphatic transport increased Aβ and tau clearance by approximately 9%–10% during a single 4h 15min sleep opportunity. Bayesian mediation analysis demonstrated that increasing EEG slow waves and declining parenchymal resistance were key mediators, and cerebrovascular compliance was a moderator, of the effect of ACX-02 on plasma AD biomarker dynamics. These findings demonstrate that pharmacologic enhancement of glymphatic transport increased brain-to-blood clearance of Aβ and tau in human participants. This suggests that enhancement of Aβ and tau clearance may serve as a complementary approach to existing disease-modifying therapies, and as a therapeutic approach in AD and AD-related proteinopathies.

## INTRODUCTION

Alzheimer’s disease (AD) is characterized by the progressive mis-aggregation of amyloid β (Aβ) and tau that begins decades before clinical symptoms of cognitive and functional impairment emerge^1,2^. Pulse-chase pharmacokinetic studies suggest that impaired clearance of Aβ was observed in the setting of aging and late-onset AD^3,4^, suggesting that impaired clearance of Aβ and tau may underlie the development of the most common sporadic form of the disease. Monoclonal antibodies targeting Aβ reduced plaque burden yet their limited effect on downstream tau pathology, long-term disease progression, and safety profile, particularly among APOEε4 carriers^5,6^, highlight the need for complementary approaches that restore endogenous clearance mechanisms.

The glymphatic system is a brain-wide perivascular network that mediates cerebrospinal fluid (CSF)-interstitial fluid (ISF) exchange and plays a central role in the removal of metabolic wastes and interstitial proteins from the brain^7–10^. In rodents, glymphatic transport facilitates clearance of Aβ and tau, is markedly enhanced during sleep, and is suppressed by sleep deprivation^7,10–13^. Neuroimaging studies in humans have confirmed the presence of perivascular glymphatic exchange in the brain^14,15^, have demonstrated that glymphatic exchange is sleep-active^16,17^, and have confirmed its role in the sleep-active brain-to-blood clearance of Aβ and tau^18^. Studies to date have yet to demonstrate that glymphatic function can be pharmacologically augmented in humans and that enhancement of glymphatic transport improves the brain-to-blood clearance of Aβ and tau. Furthermore, identifying pharmacologic targets that affect brain glymphatic transport may contribute to a more detailed understanding of the physiologic processes that regulate glymphatic transport and lead to additional therapeutic modalities to treat AD and AD-related dementias.

Central noradrenergic tone arising from projections from the locus coeruleus (LC) regulates sleep-wake states, cerebrovascular function, and brain extracellular volume fraction^10,19,20^; each of which are key determinants of glymphatic transport in rodents^10,19,21^ and humans^16–18^. Preclinical rodent studies demonstrated that broad-spectrum noradrenergic receptor blockade enhanced glymphatic function^10,22^, as did suppression of LC activity using α2-adrenergic agonists such as xylazine or dexmedetomidine (DEX)^21,23^. These observations suggest that modulation of noradrenergic tone during sleep may enhance the brain-to-blood clearance of Aβ and tau in humans.

We conducted two cross-over trials in healthy older adults to test the hypothesis that centrally acting α2A-adrenergic agonists could increase glymphatic transport and increase the brain-to-blood clearance of Aβ and tau in humans. In the first trial, DEX, a centrally acting α2A-adrenergic agonist, enhanced EEG slow-wave activity and induced systemic hypotension, but failed to increase plasma mass-balance indices of Aβ and tau clearance. We hypothesized that cerebrovascular autoregulatory vasodilation in response to systemic hypotension increased cerebral blood volume and narrowed the perivascular transport pathway, limiting any positive DEX-induced increase in glymphatic CSF-ISF exchange. To overcome this, a second trial was conducted to test ACX-02^24^, a fixed-dose combination of DEX with midodrine, a peripherally acting α1-adrenergic agonist, designed to prevent the decrease in arterial blood pressure as a consequence of central noradrenergic suppression. ACX-02 was found to increase EEG slow waves, increase cerebrovascular compliance, and reduce brain parenchymal resistance to fluid flow, thereby increasing brain-to-blood clearance of Aβ and tau.

## METHODS

### Clinical trial design

All studies were performed between December 2023 – September 2024 and were reviewed and approved by the Western Institutional Review Board (IRB No. 20232285). The studies have been registered at ClinicalTrials.gov (NCT07432997). Written informed consent was obtained from all study participants during a screening visit, prior to any study activities. Studies were carried out in accordance with the principles of the Belmont Report. The *Dexmedetomidine Trial* enrolled 9 healthy participants 56-64 years of age. The *ACX-02 Trial* enrolled 22 healthy participants 55-64 years of age.

Participants were excluded if they had cognitive impairment or clinical depression. Cognitive impairment was assessed using the Montreal Cognitive Assessment and depression was evaluated using the 15-item Geriatric Depression Scale. Participants with a self-reported history of diabetes, hypertension, coronary artery disease, pulmonary disease, neurological disease, depression, or anxiety were excluded from the study. Exclusion also applied to participants with a formal diagnosis of any sleep disorder (e.g., sleep apnea requiring positive airway pressure therapy, insomnia, restless leg syndrome, circadian rhythm sleep disorder, or parasomnia).

Participants were excluded if they had taken any prescribed or over-the-counter stimulants, sleep aids, or psychiatric medications, including antidepressants, within the past 30 days. Individuals who consumed more than 400 mg of caffeine per day, female participants who consumed more than 3 alcoholic drinks on any day or more than 7 drinks per week, and male participants who consumed more than 4 drinks on any day or more than 14 drinks per week were excluded. During intake, participants were further instructed to abstain from any sleep-altering substances throughout the study period and to avoid caffeine consumption after 12:00 p.m. on the day of the study visit. Compliance with both instructions was confirmed upon arrival at the study visit.

Two cross-over clinical trials were conducted. In the first trial, participants underwent treatment with dexmedetomidine (DEX) in the first study visit followed by placebo in the second study visit, with the two visits separated by two or more weeks (**Figure 1A**). The second trial was similar but used ACX-02 in the treatment visit. The *Dexmedetomidine Trial* was conducted at Stanford University’s Clinical Translation Research Unit (CTRU). The *ACX-02 Trial* was carried out at both the Stanford CTRU and Washington State University’s Sleep and Performance Research Center in the Human Sleep and Cognition Laboratory (**Figure 1B**). Study participants were admitted to a monitoring center at 7:00 pm on the day prior to each study visit and were required to stay awake overnight, monitored by trained research staff. On the morning of each study visit, after all-night sleep deprivation the interventions were administered during the expected morning wake period. Participants were instrumented for monitoring by electrocardiography (ECG) telemetry, percutaneous oxygen saturation (SpO_2_), nasal canula for administration of supplemental oxygen at 2 liters per minute (LPM) and continuous end-tidal CO_2_ (EtCO_2_) monitoring, and a 20 gauge (g) radial arterial catheter at the wrist for continuous systemic blood pressure measurement and blood sampling (Philips IntelliVue MP50 Patient Monitor). In addition, a 22 g intravenous catheter for drug or normal saline placebo infusion was inserted.

**Figure 1:**
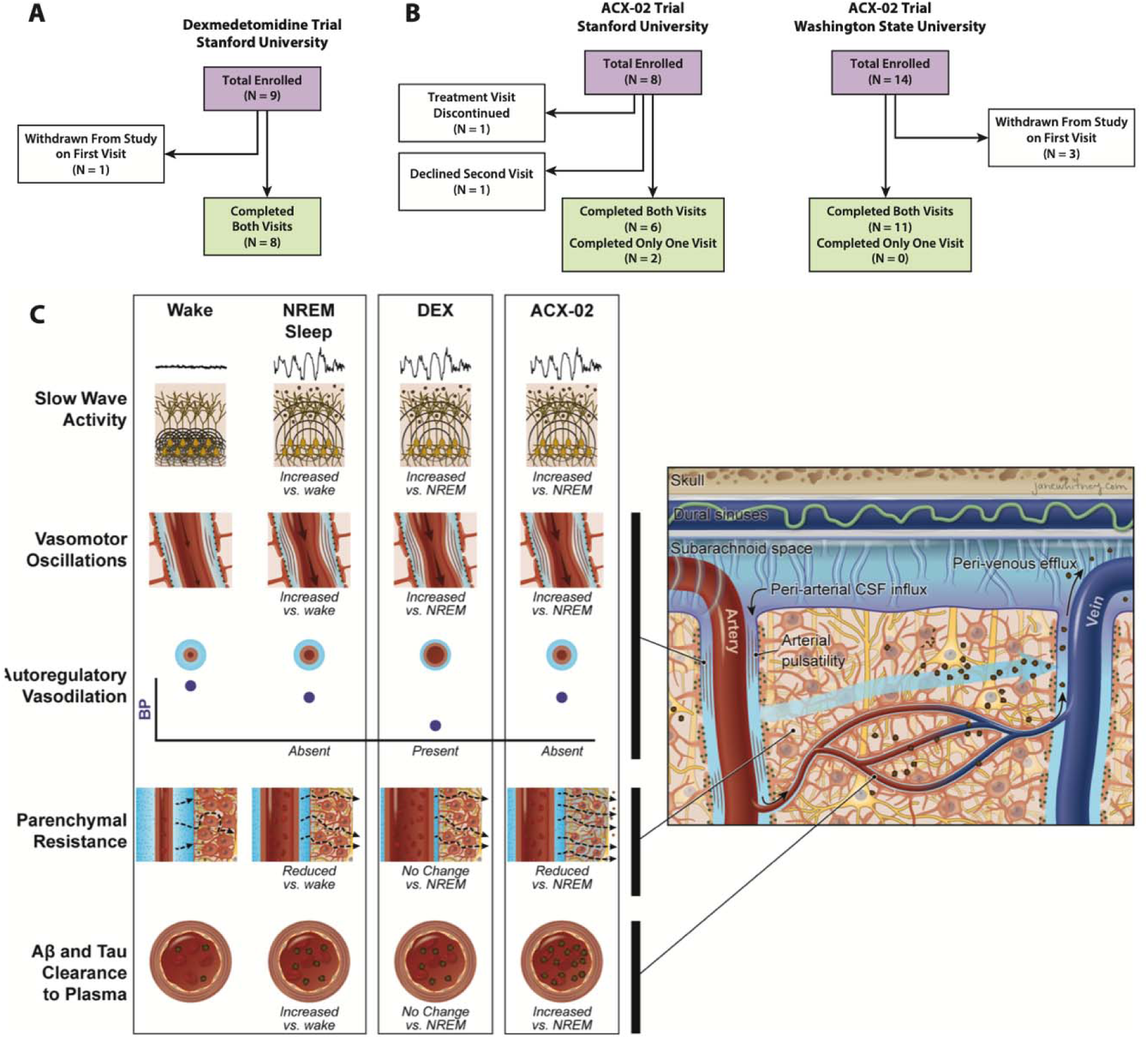
CONSORT flow diagram of participants and biological schematic. The *Dexmedetomidine Trial* was conducted at Stanford University’s Clinical and Translational Research Unit (CTRU), while the ACX-02 Clinical Trial was conducted at both Stanford University’s CTRU and Washington State University’s Sleep and Performance Research Center. Both trials used a cross-over design - treatment followed by placebo - to assess the effect of DEX on the glymphatic clearance of Aβ and tau from the brain to the blood. **(A)** The *Dexmedetomidine Trial* enrolled nine participants, of whom eight completed both study visits. **(B)** The *ACX-02 Trial* enrolled 22 participants, eight at Stanford University and 14 at Washington State University. Of the eight participants at Stanford, six completed both study visits. At Washington State University, 11 of 14 participants completed both study visits. **(C)** Illustration of the determinants of glymphatic clearance of Aβ and tau during wake, NREM sleep, DEX and ACX-02 treatment. To capture these physiological determinants, participants were instrumented with ECG telemetry, percutaneous oxygen saturation (SpO₂), nasal cannula for low-flow oxygen and continuous end-tidal CO₂ (EtCO₂) monitoring, and a radial arterial line for continuous blood pressure monitoring and blood sampling (Philips IntelliVue MP50). An intravenous line was placed for drug or saline placebo infusion. Participants were also fitted with an investigational in-ear wearable device from Applied Cognition¹L that measured key determinants of glymphatic function, including sleep features (hypnogram and spectral band power) by EEG, heart rate variability (HRV) by photoplethysmography (PPG), cerebrovascular pulse transit time (PTT*_cereb_*) by impedance plethysmography (IPG), and brain parenchymal resistance (R_P_) by dynamic electrical impedance spectroscopy.

An investigational in-ear wearable device from Applied Cognition^17^ was applied to each subject to measure key determinants of glymphatic function, including sleep features (hypnogram and spectral band power) by EEG, heart rate variability (HRV) by photoplethysmography (PPG), cerebrovascular pulse transit time (PTT*_cereb_*) by impedance plethysmography (IPG), and brain parenchymal resistance (R_P_) by dynamic electrical impedance spectroscopy (EIS), (**Figure 1C**). The Applied Cognition investigational device acquired time-multiplexed data by alternating between EEG and EIS measurements. Each acquisition cycle spanned 280 s, of which 150 s (five 30 s epochs) comprised continuous EEG recording. Reported hypnogram sleep stage durations were extrapolated to the full recording interval by scaling EEG-derived measures by a factor of 280/150, without altering any statistical conclusions. A detailed description of this device, the validation of its sleep EEG measures against gold-standard overnight polysomnography, and of the measures of R_P_ against contrast-enhanced magnetic resonance imaging (CE-MRI)-based measures of glymphatic function in humans has been previously reported^17^. The details of EEG processing, as well as hypnogram and spectral band power computations, are provided in **Supplementary Methods and Results** *Signal Processing – Electroencephalography*. Cerebrovascular PTT measurements obtained by the device using IPG were previously validated against contrast-enhanced MRI-based measures of cerebrovascular function in humans^18^.

For the *Dexmedetomidine Trial*, once instrumented, beginning at 9:00 am, participants remained supine and quietly awake for a 30-minute pre-intervention period during which baseline measurements were recorded. Following this baseline period, participants were given a 4-hour sleep opportunity during which they received a continuous intravenous (iv) infusion of DEX or placebo (normal saline, 10 ml/h). The DEX infusion was started at 0.6 μg/kg/h iv and titrated to maintain mean arterial pressure (MAP) ≥ 60 mm Hg, heart rate (HR) ≥ 45 bpm and SpO2 ≥ 92%.

For the *ACX-02 Trial*, once instrumented, beginning at 9:00 am, participants remained supine and quietly awake for a 15-minute pre-intervention period during which baseline measurements were recorded. Following this baseline period, participants were given a 4-hour and 15-minute sleep opportunity during which they received a continuous infusion of DEX iv and a single oral dose of midodrine 10 mg in the first study vist or a continuous infusion of placebo (normal saline, 10 ml/hiv) and a placebo pill in the second study visit. Midodrine or the placebo pill was administered 45 minutes before the start of the DEX or placebo iv infusion. In the *ACX-02 Trial* the DEX infusion was maintained at 0.7 μg/kg/h iv throughout the sleep period.

A starting iv bolus dose of DEX was not administered in either trial. Participant DEX iv plasma concentrations are shown in **Supplemental Figure 1**. At the end of the sleep opportunity, participants were awakened and kept supine and quietly awake for a 30-minute post-intervention measurement period.

Participants underwent peripheral blood sampling immediately prior to DEX iv infusion start, at 2 hours after the start of the infusion, immediately at the end of the DEX iv infusion, and at 30 and 60 minutes after the end of the iv infusion. These blood samples were processed and submitted for assessment of plasma DEX (iC42 at the University of Colorado Denver School of Medicine). Blood samples obtained immediately before and after the sleep intervention period underwent plasma AD biomarker assessment, including Ab40, Ab42, non-phosphorylated tau217 (np-tau217) and phosphorylated tau217 (p-tau217), using C_2_N Diagnostics’ immunoprecipitation liquid chromatography-tandem mass spectrometry platforms^25–27^. Details of specimen collection and processing are provided in **Supplementary Methods and Results** *Plasma AD biomarker assessment*.

### Change-in-baseline measures of variables

For both the *Dexmedetomidine Trial* and the *ACX-02 Trial*, the order of treatment and control visits in the trials was fixed (treatment followed by placebo) for all participants. This decision was made *a priori* to minimize patient exposure to invasive placebo administration and monitoring, particularly for participants who might poorly tolerate the drug and withdraw before the second study visit. Given both DEX and midodrine’s 2- to 3-hour elimination half-life in healthy adults^28^, and a 2- to 4-week washout period between visits, carryover effects into the placebo visit were considered to be negligible. To eliminate confounding by period drift, non-EEG estimates and plasma biomarkers were normalized to the 15-minute pre-intervention baseline immediately preceding the intervention at both the subject and visit levels. This ensured that each visit’s outcome was compared to its own preceding baseline, accounting for period drift, acclimation, and day-to-day variability. EEG power bands were compared separately during rapid eye movement (REM) and and non-REM (NREM) sleep across treatment and placebo conditions for each participant. Additionally, standardizing the timing of the sleep period controlled for confounds related to circadian influences.

### Using plasma Aβ42/Aβ40 ratio and %p-tau217 as an indicator of glymphatic clearance

In our recent pharmacokinetic and experimental study^18^, we used dynamic changes in plasma AD biomarker levels to define the effect of sleep-active glymphatic transport on the clearance of Aβ42, Aβ40, p-tau217 and np-tau217 from the brain to the blood. A key outcome of this study that was predicted by pharmacokinetic modeling and confirmed by experimental observations, was that aggregation-prone species (Aβ42 and p-tau217) and less aggregation prone species (Aβ40 and np-tau217)^29,30^ respond differentially to acute changes in glymphatic transport.

Specifically, the ratios of plasma Aβ species (Aβ42/Aβ40) and tau species (%p-tau or p-tau/np-tau) increase in response to reduced production or enhanced glymphatic clearance and decrease when production increases or clearance is impaired. This pattern reflects the dual role of non-monomeric aggregates, serving as a secondary source of monomeric Aβ42 and p-tau when production is low, and as a sink that sequesters these species when clearance is reduced^18,31^. Thus, unlike raw Aβ and tau species concentrations in plasma and CSF, which may reflect steady-state pathological burden but which cannot independently distinguish between increased glymphatic clearance and increased solute release relative to baseline, dynamic changes in plasma Aβ42/Aβ40 or %p-tau ratios enable us to infer the primary mechanism driving changes in plasma AD biomarker levels over short timescales under treatment versus placebo conditions.

### Measuring mass balance clearance of Aβ42/Aβ40 and %p-tau217

Plasma AD biomarkers represent concentrations in plasma, and treatment-induced changes in these concentrations and their ratios can be confounded by fluctuations in plasma volume during the intervention period. To address this, the post- to pre-intervention change in the Aβ42/Aβ40 and %p-tau217 mass ratios were analyzed, that provided a mass balance measure of preferential clearance of Aβ42 relative to Aβ40, and p-tau relative to np-tau, from the ISF to plasma during 4 hor 4 h15 min sleep opportunity under treatment versus placebo conditions. As shown in the **Supplementary Methods and Results** *Derivation of mass balance ratios*, the ratio of plasma Aβ42/Aβ40 concentrations post-versus pre-intervention directly reflects the change in total plasma mass of Aβ42 relative to Aβ40. Crucially, this ratio is unaffected by changes in plasma volume over the intervention period. The same principle applies to %p-tau217.

### Development of a linear mixed models to define the effect of drug interventions on plasma AD biomarkers and determinants of glymphatic function

We developed a series of linear mixed models to assess the treatment effects of DEX and ACX-02 on the clearance of Aβ and tau from the brain to the plasma. Our primary outcomes of interest were the individual treatment effects on Aβ and tau. In these models, the dependent variables were the post-intervention versus pre-intervention ratios of plasma Aβ42/Aβ40 and %p-tau217, analyzed separately for Aβ and tau. Potential confounding variables age, sex, APOE-ε4 status, and study site were included as fixed effect covariates. Participant ID was modeled as a random intercept.

Because the *Dexmedetomidine Trial* was conducted across a range of intravenous infusion rates, titrated to maintain MAP and HR above predefined minimum thresholds, different participants experienced variable levels of total DEX exposure during the treatment period (**Supplemental Figure 1**). We used this variability to develop a linear mixed model relating total systemic DEX exposure under treatment conditions to the observed Aβ and tau ratios. Total DEX exposure was calculated using the trapezoidal rule to estimate the area under the curve (AUC) based on plasma concentrations measured at infusion start, after 2 hours and at infusion stop. A multivariate linear mixed model was then constructed with the Aβ and tau ratios as dependent variables. This model included both a random intercept for participant as well as age, sex, and APOE-ε4 status as potential confounding effect covariates.

A series of linear mixed models was next developed to assess the pharmacodynamic effects of DEX and ACX-02 on potential key mediators and suppressors of glymphatic function. For all non-EEG measures, the dependent variables were the mean of the values over the treatment period normalized to the mean of the values over the15 min pre-treatment baseline. For EEG data, relative power within frequency bands was calculated by normalizing to total EEG power (0.5 Hz–50 Hz) and compared separately during REM and NREM sleep across treatment and placebo conditions for each participant. Potential confounding variables age, sex, APOE-ε4 status, and study site were included as fixed effect covariates. Participant ID was modeled as a random intercept.

### Development of a Bayesian multivariate linear mixed mediation model identifying pharmacodynamic mediators and suppressors of glymphatic clearance

We developed a series of Bayesian multivariate linear mixed models to identify the mediators and suppressors of the effect of ACX-02 on the glymphatic clearance of Aβ and tau to plasma, using the *brms* package in R. The joint model tested the full set of selected candidates as potential mediators and suppressors. These candidates were drawn from eight physiological categories based on published studies defining glymphatic function in rodents and humans^10,17–19,21,32,33^: 1) Sleep-associated modulation of extracellular volume, 2) Cerebral vasomotor oscillations, 3) Peripheral vascular tone, 4) EEG slow wave oscillations, 5) EEG NREM and REM power bands, 6) EEG-derived hypnographic sleep stage durations, 7) Sympathetic and parasympathetic balance, 8) respiratory measures.

Within each category, individual candidate factors were included in the mediation analyses if their effect size under ACX-02 treatment exceeded 0.5 SD. This selection process yielded the following variables for the joint mediation analysis:

#### Sleep-associated modulation of extracellular volume

Parenchymal resistance (R_P_) met the selection criteria.

#### Cerebral vasomotor oscillations

Cerebrovascular pulse transit time (PTT*_cereb_*), a measure of pulsatility or oscillation amplitude, met the selection criteria.

#### Slow waves

Slow wave count met the selection criteria.

#### Peripheral vascular tone

Three candidates were initially considered: (1) peripheral pulse transit time (PTT*_periph_*), measured as the time delay from the electrocardiogram R wave to the peak of the radial arterial line pressure pulse; (2) HR during NREM sleep; and (3) mean arterial pressure (MAP) during NREM sleep. MAP during NREM sleep was not statistically significant between ACX-02 and placebo, and PTT*_periph_*and HR during NREM sleep were strongly correlated (r = –0.60), making them too collinear to include together. Therefore, only PTT*_periph_*was selected.

#### EEG power bands

Due to high collinearity between power bands within and across NREM and REM sleep, only three bands were selected based on consistency with prior rodent and human studies of glymphatic function: NREM slow delta (0.5–1 Hz), NREM delta (1–4 Hz), and REM beta (15–30 Hz).

#### Hypnogram sleep stage durations

Only N2 and wakefulness after sleep onset (WASO) sleep stage durations met the effect size inclusion criteria and were selected.

#### Autonomic nervous system measures

Neither the standard deviation of normal-to-normal intervals (SDNN heart rate variability) nor the low-frequency to high-frequency HRV met the significance threshold.

#### Respiratory measures

Neither respiratory rate nor EtCO_2_ met the criteria for inclusion.

Parenchymal Resistance and PTT*_cereb_*had interaction effects with ACX-02 treatment and were each included as mediators and moderators in the Bayesian model by including interaction terms. The selected variables from the three candidate mediator groups - *Slow waves*, *EEG power bands*, and *Hypnogram sleep stage durations* - were strongly correlated with one another, with correlation coefficients ranging from 0.44 to 0.76. This level of collinearity made it unsuitable to include all variables in a single Bayesian model. Therefore, three separate Bayesian multivariate linear mixed models were constructed, each identical in structure except for the inclusion of one of the three preceding variable groups.

All selected mediator and suppressor variables were normalized to have a mean of zero and a standard deviation of one. The treatment variable was centered by coding −0.5 for placebo and 0.5 for ACX-02 treatment. The Bayesian models used default weakly informative priors.

The models assessing the effect of ACX-02 on mediator variables were Bayesian versions of the linear mixed models used in prior pharmacodynamic analyses^18^. These included age, sex, APOEε4 status, and study site as fixed effect covariates, and participant ID as a random intercept.

The plasma Aβ and tau outcome model of the mediation analysis was a Bayesian multivariate model that included the full set of mediators and suppressors, a biomarker indicator variable, and the same fixed effect covariates: age, sex, apolipoprotein APOEε4 status, and study site. Participant ID was modeled as a random intercept, and a biomarker-specific random slope was added to account for the multivariate structure of the Aβ and tau dependent variables.

## RESULTS

Statistical significance tests reported herein were not adjusted for multiple comparisons. With the exception of hypnogram measures, physiological variables were Winsorized at the 2.5th and 97.5th percentiles to mitigate the influence of potential outliers.

### Study participant information

A Consolidated Standards of Reporting Trials (CONSORT) diagram for the *Dexmedetomidine Trial* and *ACX-02 Trial* is shown in **Figure 1A-B**. In the *Dexmedetomidine Trial*, nine participants were enrolled, of whom eight completed both study visits (61.2 ± 3.3 years of age; 4 female).

One participant was withdrawn by the site investigator during the first study visit due to non-compliance with protocol instructions. The *ACX-02 Trial* enrolled 22 participants, eight at Stanford University and 14 at Washington State University, of which 19 completed at least one study visit (60.6 ± 3.2 years of age; 12 female). At Stanford, six of the eight participants completed both study visits and two participants completed one study visit. Of the latter two, one did not return for the second study visit, while the other was restless during the second study visit sleep period, repeatedly sitting up, resulting in the early termination of the study visit. At Washington State University, 11 of the 14 participants completed both study visits. Three were withdrawn after the first study visit due to inadequate DEX delivery by the infusion pump, likely resulting from a change in anesthesiology staff midway through the trial. Plasma tau data from one of the remaining participants were excluded from analysis because pre-intervention p-tau217 concentrations were below the lower limit of quantification (0.65 pg/mL) at both placebo and treatment visits, while post-intervention concentrations were markedly elevated (2.93 and 3.94 pg/mL, respectively).

Participant demographics, as well as MoCA and GDS scores, are presented in **Table 1**. No significant differences were observed between the two trials for any of these measures. The average MoCA scores for the *Dexmedetomidine Trial* and *ACX-02 Trial* were 25.6L±L3.2 and 26.6L±L2.7, respectively, both falling within age-normative ranges^34^.

**Table 1:** Baseline characteristics of enrolled participants in the *Dexmedetomidine* and *ACX-02 Trials*\*.

|  | **Dexmedetomidine**<br>**Trial** | **ACX-02 Trial** | **P value** |
| --- | --- | --- | --- |
| **Number** | 8 | 19 |  |
| **Age (y)** | 61.2 ± 3.3 | 60.6 ± 3.2 | 0.662 |
| **Female Sex** | 4 (50.0%) | 12 (63.2%) | 0.675 |
| **APOE-ε4 Positive** | 1 (12.5%) | 6 (31.6%) | 0.633 |
| **Amyloid Plaque Positive** | 1 (12.5%) | 3 (15.8%) | 1.00 |
| **MoCA** | 25.6 ± 3.2 | 26.6 ± 2.7 | 0.469 |
| **GDS** | 0.2 ± 0.5 | 0.8 ± 1.2 | 0.0788 |

\*Values are Mean ± SD or count (%). P-values are derived from Student's t-test for Age, MoCA, and GDS, and from Chi-square test (or Fisher's exact test) for the other characteristics. MoCA, Montreal Cognitive Assessment; GDS, Geriatric Depression Scale. Amyloid plaque status determined using C<sub>2</sub>N PrecivityAD test with cutoff of 0.089<sup>48</sup>.

### The effect of DEX on glymphatic function and sleep-related Ab and tau clearance

Summary plasma AD biomarker levels for the *Dexmedetomidine Trial* and *ACX-02 Trial* pre-and post-intervention of placebo or drug are provided in **Supplemental Table 1**. Plasma levels and treatment-to-control ratios for Ab40, Ab42, Ab42/Ab40, np-tau217 and %p-tau217 are shown. Only two participants (one in the *Dexmedetomidine Trial* and one in the *ACX-02 Trial*) had p-tau217 values that did not fall below the limit of detection (LOD) of 1.3 pg/ml and p-tau217 was excluded from the table. For samples in which the p-tau217 concentration was below the limit of detection (LOD), the %p-tau217 was calculated using an imputed p-tau217 value equal to half the LOD^25^.

Our prior study demonstrated that dynamic changes in plasma Aβ42/Aβ40 or p-Tau/np-Tau ratio are consistent with decreased production or increased clearance of Aβ and tau biomarkers^18^. Our initial linear mixed effects model demonstrated that DEX treatment showed no treatment (drug versus placebo) effect on either plasma AD biomarker ratio (**Table 2**). We next assessed whether DEX had an exposure-response effect on plasma AD biomarker ratios. The area under the curve (AUC) for plasma concentration was calculated over the 4-hour intravenous infusion, based on DEX plasma concentrations at infusion start, at 2 hours, and at the end of the infusion. The two AD biomarker ratios were modeled jointly using a linear mixed-effects model applied to stacked data, with a biomarker indicator predictor and a participant-level random intercept to account for within-subject correlation. The effect of DEX AUC on the plasma AD biomarker ratios from pre-treatment to post-treatment showed that increasing DEX administration increased the Aβ42/Aβ40 ratio (estimate 0.352 ng·h/mL; 95% CI: 0.104–0.600 ng·hr/mL; P = 0.008) after adjusting for age, sex, and APOE ε4 status. The association was significantly stronger for %p-tau217, with an additional effect of 0.189 relative to Aβ42/Aβ40 (p = 0.031), corresponding to an estimated effect size of 0.541 (**Table 3**).

**Table 2:** Effect of DEX treatment versus placebo on plasma biomarker ratios Aβ42/Aβ40 and %p-tau217*.

| Predictor Group | Predictor | | Aβ42/Aβ40 | | | | %p-tau217 | | | |
| --- | --- | --- | --- | --- | --- | --- | --- | --- | --- | --- |
|  |  |  | Estimate | DF | CI | P | Estimate | DF | CI | P |
| Fixed Effects | Age |  | -0.003 | 8 | -0.015 – 0.009 | 0.618 | -0.026 | 16 | -0.068 – 0.017 | 0.215 |
|  | Sex (male) |  | 0.016 | 8 | -0.065 – 0.097 | 0.656 | 0.208 | 16 | -0.075 – 0.492 | 0.139 |
|  | APOE-e4 (positive) |  | 0.090 | 8 | -0.033 – 0.213 | 0.129 | -0.023 | 16 | -0.454 – 0.407 | 0.910 |
|  | Predictor Effects | Treatment | 0.043 | 8 | -0.031 – 0.118 | 0.217 | 0.218 | 16 | -0.044 – 0.480 | 0.097 |
| Random Effects | $\sigma^2$ | | 0.004 | | | | 0.061 | | | |
| | $\tau_{00pid}$ | | 0.000 | | | | 0.000 | | | |

\*DF, Satterthwaite degrees of freedom; CI, 95% confidence interval; P, p-value; $\sigma^2$ , residual variance; $\tau_{00}$ , between subject intercept variance. Two-sided *p*-values were calculated using a *t*-distribution with Satterthwaite degrees of freedom and were not adjusted for multiple comparisons. **Bolded P values reflect significant associations at P < 0.05.** Shaded cells demark random effects and potential biological confounders.

**Table 3:** Changes in plasma Aβ42/Aβ40 and %p-tau217 with changes in DEX AUC exposure*.

| Predictor Group | Predictor |  | Estimate | DF | CI | P |
| --- | --- | --- | --- | --- | --- | --- |
| Fixed Effects | Age |  | -0.072 | 16 | -0.108 – -0.036 | **0.001** |
|  | Sex (male) |  | 0.422 | 16 | 0.177 – 0.668 | **0.002** |
|  | APOE-e4 (positive) |  | 0.355 | 16 | -0.011 – 0.699 | **0.044** |
|  | Biomarker Identity (%p-tau217) |  | 0.189 | 16 | 0.020– 0.358 | **0.031** |
|  | Predictor Effects | DEX AUC [ng·h/mL] | 0.352 | 16 | 0.104 – 0.600 | **0.008** |
| Random Effects | $\sigma^2$ | | 0.025 | | | |
| | $\tau_{00pid}$ | | 0.000 | | | |

\*DF, Satterthwaite degrees of freedom; CI, 95% confidence interval; P, p-value; $\sigma^2$ , residual variance; $\tau_{00}$ , between subject intercept variance; AUC, area under the curve calculated over the 4-h intervention period. The Biomarker Identity predictor is a fixed-effect indicator with value 0 for Aβ42/Aβ40 and 1 for %p-tau217. Two-sided *p*-values were calculated using a *t*-distribution with Satterthwaite degrees of freedom and were not adjusted for multiple comparisons. **Bolded P values reflect significant associations at P < 0.05.** Shaded cells demark random effects and potential biological confounders.

To explain the absence of a treatment effect despite a significant exposure-response relationship, we examined DEX’s influence on pharmacodynamic mediators and suppressors of glymphatic clearance (**Supplemental Table 2**). DEX increased slow wave count (placebo: 202.1 ± 196.8; treatment: 406.6 ± 338.1; P = 0.007), as well as power in the NREM EEG slow delta and delta bands (P < 0.001 and P = 0.025, respectively), and decreased power in the REM EEG beta band (P = 0.001). These changes (**Figure 2A**) in sleep microstructure mediate glymphatic clearance in rodents^21^ and in human participants^17^. However, DEX did not decrease parenchymal resistance R_P_ (**Supplemental Table 2** and **Figure 2B**), a key mediator of glymphatic clearance^10,17^. This is in contrast to findings from rodent studies using the α2 agonist xylazine, which increased brain extracellular volume fraction. It is possible that other competing effects of DEX account for this discrepancy.

**Figure 2:**
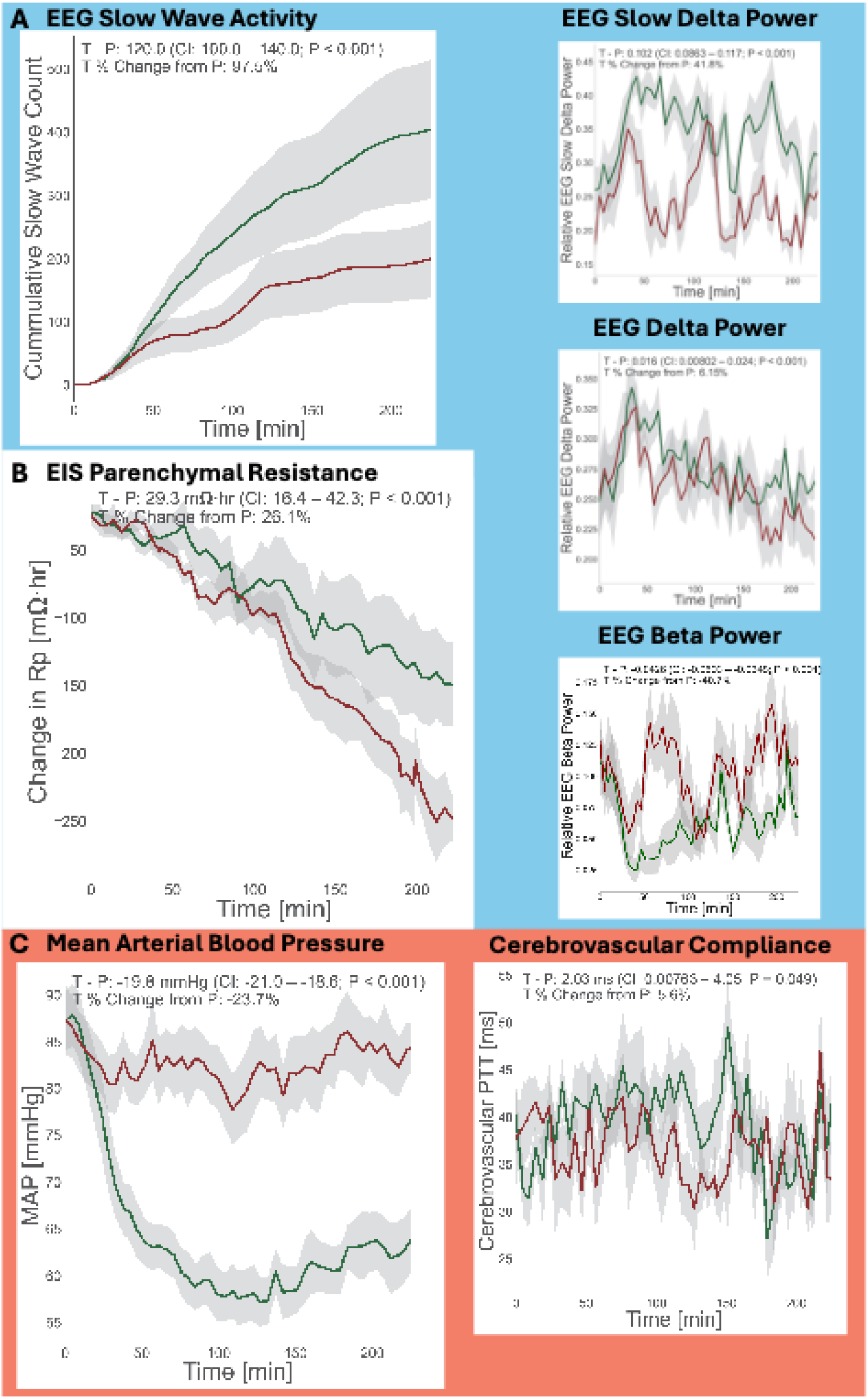
DEX treatment effect on potential mediators of glymphatic clearance. The effect of DEX (green) versus placebo (red) in the *Dexmedetomidine Trial* over the 4-hour sleep opportunity are shown for **(A)** slow wave count, NREM EEG slow delta bandpower (0.5Hz – 1Hz), and REM EEG beta power (15Hz – 30Hz); **(B)** parenchymal resistance (R_P_); and **(C)** mean arterial pressure (MAP) and cerebrovascular compliance (PTT*_cereb_*). DEX induced a 5.6% increase in PTT*_cereb_*(P = 0.049) that is associated with an increase in cerebral blood volume^18^. Plotted data consists of measured values recorded every 280 seconds, with linear interpolation applied to achieve a one-minute resolution. Standard error of the mean for each plot is shown in light grey. The reported statistical results include the treatment effect size, 95% confidence interval (CI) and two-sided p-values for fixed effects were obtained using Wald t-tests with Satterthwaite degrees of freedom from a linear mixed-effects model fitted to the measured values. This model includes a random intercept for participant ID and is adjusted for age, sex, and APOEε4 status. The reported percent change reflects the treatment effect size as a percentage of the marginal placebo mean. P values were not adjusted for multiple comparisons. Placebo (P) and Treatment (T).

DEX treatment significantly decreased NREM MAP (placebo: 0.910 ± 0.410; treatment: −0.796 ± 0.551; P < 0.001; in units of SD). Observing changes in MAP throughout the sleep period, MAP exhibited a marked drop with DEX early in the 4-hour sleep period, which was sustained throughout the duration of the period. The average MAP decrease over the sleep period was 19.8 mmHg (95% CI: −21.0 – −18.6; P < 0.001; **Figure 2C**). This reduction in MAP was associated with a 2.03Lms (95% CI: 0.0076 – 4.05; P = 0.049) increase in PTT*_cereb_*, representing a 5.6% increase compared to placebo (**Figure 2C**). In our prior study, dynamic changes in PTT*_cereb_* were associated with shifts in cerebral blood volume (CBV), with longer PTT values aligning with increased CBV^18^. Thus, this increase in PTT*_cereb_* likely reflects autoregulatory vasodilation resulting from DEX-induced systemic hypotension. A recent study in rodents demonstrated that cerebral vasodilation resulting from hypotension suppresses glymphatic function by closing off perivascular spaces upstream of the brain interstitium^33^. Thus we propose that autoregulatory vasodilation may explain the higher measured R_P_ under DEX (**Figure 2B**) and the absence of a treatment effect of DEX on Aβ and tau ratios despite significant enhancements in slow-wave oscillation count, increased NREM EEG slow delta and delta band power, and decreased REM EEG beta band power during the DEX treatment sleep period (**Figures 2A**).

### Peripheral α1 adrenergic agonism offsets the effect of cerebrovascular autoregulation on glymphatic clearance

We next tested the mechanistic hypothesis that DEX-induced systemic hypotension triggered cerebral autoregulatory vasodilation that suppressed glymphatic clearance in the face of increased slow waves. If true, preventing hypotension or maintaining systemic arterial pressure during DEX iv administration should reduce parenchymal resistance R_P_ and improve glymphatic clearance.

Midodrine is an oral α1-adrenergic agonist with an active metabolite, desglymidodrine, which has a plasma half-life of approximately 3 to 4 hours that is comparable to DEX’s plasma half-life of 2 to 3 hours. Midodrine does not cross the blood–brain barrier, making it a peripherally acting α1-adrenergic agonist, with effects limited to the peripheral vascular system^35^. Midodrine is used to treat symptomatic orthostatic hypotension; thus we administered it prior to DEX iv infusion to prevent hypotension and maintain the arterial pressure.

We used a fixed-dose combination (ACX-02) consisting of midodrine 10 mg oral administered 45 minutes prior to the start of a DEX infusion at 0.7 μg/kg/h iv that was continued throughout a 4-hour and 15-minute sleep period. Examination of pharmacodynamic mediators and suppressors of glymphatic clearance under ACX-02 (**Supplemental Table 3** and **Figure 3**) revealed that, similar to DEX alone, this fixed-dose drug combination increased slow wave count (placebo: 427.3 ± 329.8; treatment: 764.9 ± 467.8; P < 0.001), as well as power in the NREM EEG slow delta and delta bands (both P < 0.001), and decreased power in the REM EEG beta band (P < 0.001) as shown in **Figure 3A**. In contrast to DEX alone, ACX-02 decreased R_P_ by 0.902 SD (P < 0.001) and had no significant effect on NREM MAP (P = 0.160) as shown in **Figures 3B** and **3C**.

**Figure 3:**
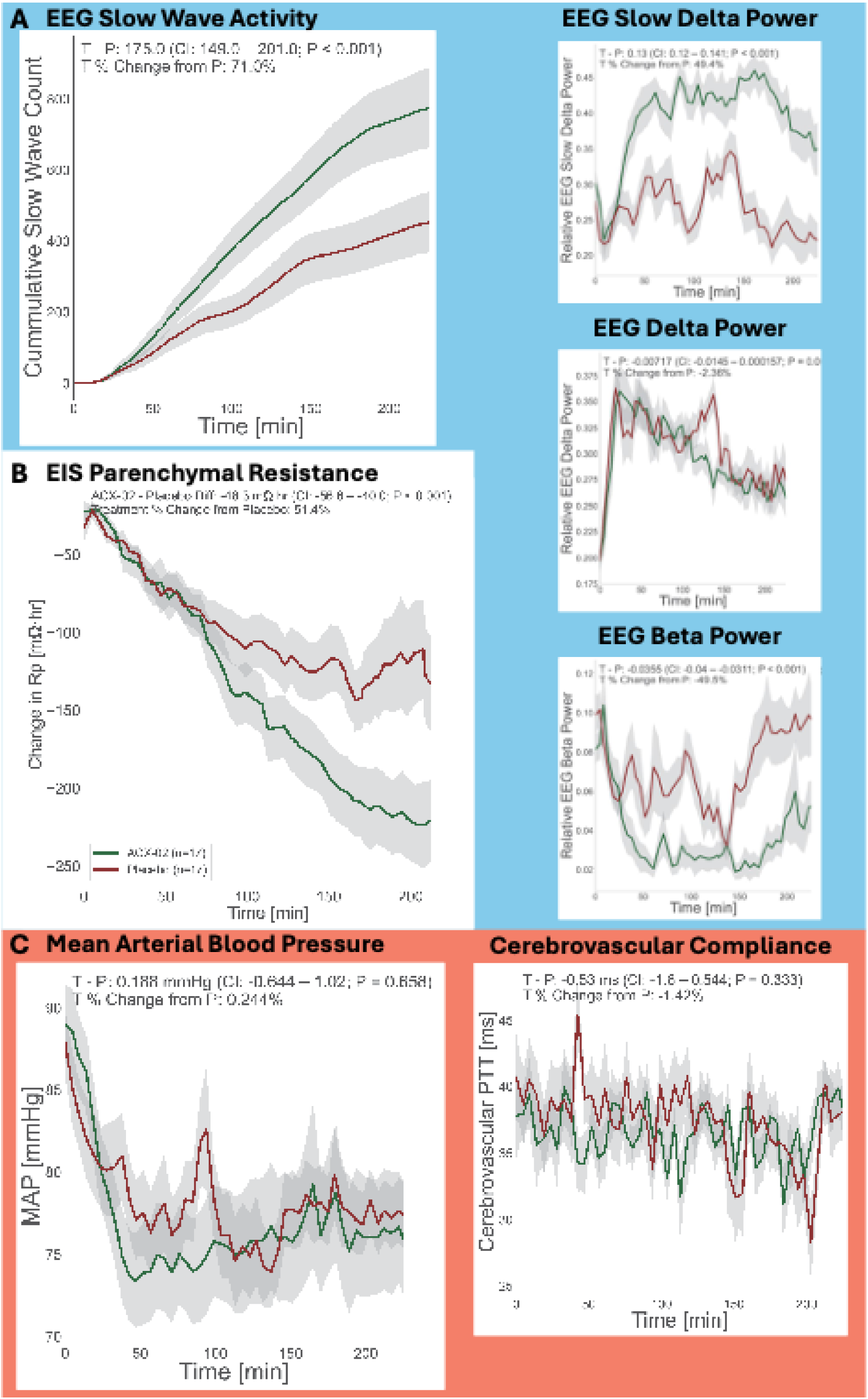
ACX-02 treatment effect on potential mediators of glymphatic clearance. The effect of ACX-02 (green) versus placebo (red) over the *ACX-02 Trial* 4-hour and 15-minutes sleep opportunity are shown for **(A)** slow wave count, NREM EEG slow delta power (0.5Hz – 1Hz), and REM EEG beta power (15Hz – 30Hz); **(B)** parenchymal resistance (R_P_); and **(C)** mean arterial pressure (MAP) and cerebrovascular compliance (PTT*_cereb_*). ACX-02 normalized vascular pressures and was not associated with an increase in PTT*_cereb_*that has been associated with an increase in cerebral blood volume^18^. Plotted data consists of measured values recorded every 280 seconds, with linear interpolation applied to achieve a one-minute resolution. Standard error of the mean for each plot is shown in light grey. The reported statistical results include the treatment effect size, 95% confidence interval (CI) and two-sided p-values for fixed effects were obtained using Wald t-tests with Satterthwaite degrees of freedom from a linear mixed-effects model fitted to the measured values. This model includes a random intercept for participant ID and is adjusted for age, sex, and APOEε4 status. The reported percent change reflects the treatment effect size as a percentage of the marginal placebo mean. P values were not adjusted for multiple comparisons. Placebo (P) and Treatment (T).

With ACX-02, the MAP recorded over the 4-hour and 15-minute sleep intervention period did not differ significantly from placebo (0.188 mmHg; 95% CI: −0.644 – 1.02; P = 0.658) (**Figure 3C**). The early approximate 10% decrease in MAP observed in the placebo group of both the *Dexmedetomidine Trial* and *ACX-02 Trial*, as well as in the ACX-02 treatment group, is consistent with known physiological reductions in blood pressure during the transition from wakefulness to sleep^36^. There was likewise no significant difference in PTT*_cereb_* between ACX-02 and placebo (−0.53 ms; 95% CI: −1.6 – 0.54; P = 0.333; **Figure 3C**), confirming that normalizing peripheral blood pressure effectively avoided the cerebral autoregulatory vasodilation response observed with DEX alone. In contrast to DEX alone, ACX-02 significantly reduced R_P_ by −48.3 mΩ·h (95% CI: −56.6 – −40.0; P < 0.001, **Figure 3B**), representing a 51.4% decrease from placebo. Similar to DEX alone, we observed significant enhancements in slow-wave oscillation count, increased NREM EEG delta power, and decreased REM EEG beta power during the ACX-02 sleep period (**Figure 3A**).

### Effects of ACX-02 on glymphatic function and sleep-related Ab and tau clearance

We next evaluated the effect of ACX-02 on brain-to-blood clearance of Ab and tau. ACX-02 treatment showed a significant effect on plasma clearance of Aβ42/Aβ40 (effect size, 0.084; 95% CI 0.012 – 0.157; P = 0.024) and of %p-tau217 (effect size, 0.108; 95% CI 0.001 – 0.215; P = 0.049) (**Table 4**). The percent increase in clearance to plasma of treatment over placebo was 8.45% for Aβ42/Aβ40 and 9.66% for %p-tau217.

**Table 4:** Effect of fixed-dose combination of DEX and midodrine ACX-02 treatment versus placebo on plasma Aβ42/Aβ40 and %p-tau217*.

| Predictor Group | Predictor | | Aβ42/Aβ40 | | | | %p-tau217 | | | |
| --- | --- | --- | --- | --- | --- | --- | --- | --- | --- | --- |
|  |  |  | Estimate | DF | CI | P | Estimate | DF | CI | P |
| Fixed Effects | Age |  | -0.001 | 36 | -0.012 – 0.011 | 0.925 | 0.024 | 19 | -0.033 – 0.081 | 0.388 |
|  | Sex (male) |  | 0.049 | 36 | -0.027 – 0.125 | 0.203 | -0.160 | 19 | -0.519 – 0.199 | 0.362 |
|  | APOE-e4 (positive) |  | 0.032 | 36 | -0.046 – 0.109 | 0.411 | 0.257 | 19 | -0.117 – 0.632 | 0.167 |
|  | Study-Site (Stanford) |  | 0.059 | 36 | -0.017 – 0.134 | 0.122 | -0.063 | 19 | -0.419 – 0.293 | 0.716 |
|  | Predictor Effects | Treatment | 0.084 | 36 | 0.012 – 0.157 | **0.024** | 0.108 | 15 | 0.001 – 0.215 | **0.049** |
| Random Effects | $\sigma^2$ | | 0.012 | | | | 0.018 | | | |
| | $\tau_{00pid}$ | | 0.000 | | | | 0.117 | | | |

\*DF, Satterthwaite degrees of freedom; CI, 95% confidence interval; P, p-value; $\sigma^2$ , residual variance; $\tau_{00}$ , between subject intercept variance. Two-sided $p$ -values were calculated using a $t$ -distribution with Satterthwaite degrees of freedom and were not adjusted for multiple comparisons. **Bolded P values reflect significant associations at P < 0.05.** Shaded cells demark random effects and potential biological confounders.

Comparisons between placebo and treatment pharmacodynamic and biomarker values averaged over the 4-hour and 15-minute sleep opportunity and normalized to the corresponding baseline are presented in **Supplemental Figure 2** for both DEX and ACX-02. Correlations between these measures under DEX alone and ACX-02 treatment are illustrated in **Supplemental Figure 3**. Key drivers of glymphatic clearance - R_P_, PTT*_cereb_*, EEG slow-wave oscillations, and EEG power bands – show distinct differences between treatments. ACX-02 produced stronger correlations among EEG power bands and R_P_, and weaker correlations with PTT*_cereb_*. These patterns align with expectations for ACX-02’s mechanism of action, suggesting it more effectively coordinates these drivers of glymphatic function to enhance clearance.

### Pharmacodynamic mediators and suppressors of ACX-02 treatment effect on AD biomarker clearance to plasma

We next sought to determine the extent to which the measured physiological determinants of glymphatic transport either mediated or suppressed the effect of ACX-02 on increasing Aβ and tau clearance to plasma. **Supplemental Table 3** lists the effect of ACX-02 on each of the eight monitored physiological parameter groups and related individual features. Applying a stringent effect size > 0.5 SD eliminated HRV (autonomic nervous system balance) and respiratory measures.

To address multicollinearity among slow wave count, EEG power bands, and sleep hypnogram measures, we developed three separate Bayesian linear mixed models – **Slow Wave**, **EEG**, and **Hypno**. These models were structurally identical, each including R_P_, PTT*_cereb_*, and PTT*_periph_*, but differed in that each incorporated only one of the three variable sets: slow wave count, EEG power bands, or hypnogram measures. Furthermore, parenchymal resistance R_P_ and PPT*_cereb_* were both included as mediators and moderators in all three models as they showed a treatment-dependent interaction effect. Bayesian mediation analysis for each model used weakly informative priors and 8,000 simulations to estimate the mean and the 95% Bayesian confidence intervals for the Average Causal Mediation Effect (ACME, or Indirect Effect).

All three models revealed that parenchymal resistance R_P_ was a strong mediator and that its mediation effect was enhanced by ACX-02 versus placebo. These models also showed that NREM PPT*_cereb_* was a suppressor in placebo sleep, likely through an increase in CBV, and that ACX-02 treatment shifted PPT*_cereb_* from a suppressor to a mediator of clearance. Thus, these two pathways showed ACX-02-dependent engagement in increasing Aβ and tau clearance to plasma acting mechanistically as mediator-moderators (**Figure 4** and **Supplemental Table 4**).

**Figure 4:**
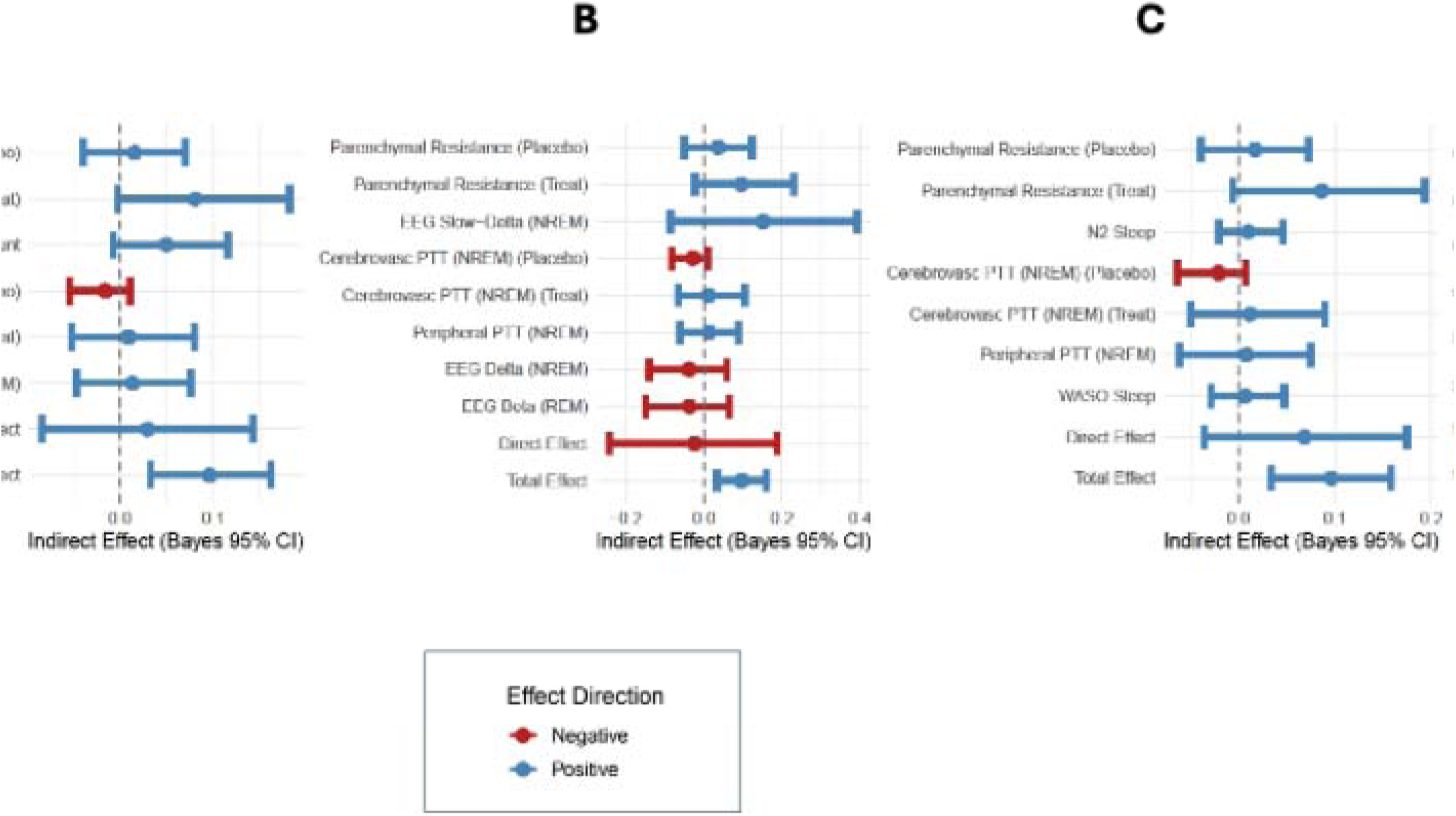
Indirect mediation effect, direct effect and total effect of ACX-02 on glymphatic clearance. Several physiological variables that were continuously monitored during the ACX-02 intervention, as compared to placebo, responded significantly to ACX-02 treatment, with standardized effect sizes exceeding 0.5. These were subsequently tested for their role in mediating or suppressing the glymphatic clearance of AD biomarkers to plasma. These variables corresponded to six key physiological domains involved in glymphatic clearance: (i) astrocyte modulation of brain interstitial resistance (Parenchymal Resistance); (ii) cerebral vasomotor oscillations (Cerebrovascular PTT (NREM)); (iii) slow waves (Slow Wave Count); (iv) EEG power bands during NREM and REM sleep (EEG Slow Delta Power (NREM), EEG Delta Power (NREM), EEG Beta Power (REM)); (v) peripheral vascular tone (Peripheral PTT (NREM)); (vi) hypnogram sleep stage duration (N2 sleep and WASO). Measures derived from autonomic nervous system balance (sympathetic/parasympathetic) and respiratory function did not show a significant response to ACX-02 and were excluded from the mediation analysis. The selected candidate mediators/suppressors were standardized (zero mean, unit variance) before conducting Bayesian mediation analysis using weakly informative priors. The three sleep-related groups of measures—slow waves, EEG power bands, and hypnogram sleep stage durations—were strongly correlated with one another. Therefore, three separate Bayesian multivariate linear mixed models were constructed, each including Parenchymal Resistance, Cerebrovascular PTT (NREM), and Peripheral PTT (NREM) and further including variables from one of: **(A)** slow waves, **(B)** EEG power bands, or **(C)** hypnogram sleep stage durations. The analysis involved 8,000 simulations to estimate the mean and the 95% Bayesian confidence intervals (Bayes 95% CI). Since ACX-02 treatment produced a positive effect on AD biomarker clearance, a positive effect direction occurred if ACX-02 either increased (or decreased) a physiological variable relative to placebo, and that same increase (or decrease) in the variable led to greater clearance of AD biomarkers. Hence a positive effect was considered mediating. Conversely, a negative effect direction occurred when the effect of ACX-02 on the physiological variable was opposite to that variable’s impact on clearance. In this case, the variable was considered to suppress the effect of ACX-02. Bayesian one-sided P values were computed from the 8,000 simulations from the posterior probability mass crossing zero.

As demonstrated above, that reductions in MAP induced a cascade of cerebrovascular changes characterized by increased CBV and prolongation of cerebrovascular pulse-transit time, PTT*_cereb_*. This hemodynamic pathway accounts for the marked increase in PTT*_cereb_*observed under DEX, which was accompanied by significant systemic hypotension (**Figure 2C**; **Supplementary Figures 2C** and **2M**). The resulting elevation in CBV and reduced vascular compliance are conditions that would be expected to reduce the perivascular space volume and impair glymphatic exchange. In contrast, ACX-02 increased PTT*_cereb_* relative to placebo in the absence of measurable changes in MAP (**Figure 3C**; **Supplementary Figures 2C** and **2M**).

Moreover, ACX-02 similarly prolonged peripheral vascular PTT (**Supplementary Figure 2L**), indicating a coordinated central and systemic modulation of vascular transit dynamics that is independent of arterial pressure.

When expressed as a fraction of the total treatment effect, R_P_ accounted for 12.2–31.3% of mediation under placebo conditions across the **Slow Wave**, **EEG**, and **Hypno** Bayesian models, increasing to 68.1–81.0% under ACX-02 treatment (**Supplementary Table 4**). This marked amplification indicates that ACX-02 substantially enhances the capacity of R_P_ to mediate glymphatic clearance, consistent with effects of ACX-02 on other physiological drivers of glymphatic transport, such as EEG slow waves and cerebrovascular dynamics. In parallel, the suppressive influence of cerebrovascular pulse transit time (PTT*_cereb_*) was attenuated by ACX-02, shifting from −23.8% to −13.1% under placebo to 8.7–10.9% under treatment across models. This reversal suggests that ACX-02 alleviates the cerebrovascular constraints that otherwise limit clearance and priming R_P_-dependent processes for more effective engagement.

Slow-wave count and EEG slow-delta power exhibited large fractional contributions (42.0% and 130.8%, respectively), with values exceeding 100% reflecting compensation for concurrent suppressive pathways. In contrast, N2 sleep duration and WASO displayed minimal mediation (8.0% and 5.8%), indicating limited involvement of sleep stage parameters in treatment-responsive clearance mechanisms. Notably, NREM EEG delta and REM EEG beta power appeared as suppressors because ACX-02 treatment shifted the EEG spectrum toward the slow delta band, reducing relative power in other bands (**Supplemental Table** 3). Peripheral pulse transit time showed negligible mediation, consistent with the central vascular specificity of ACX-02’s mechanism of action. Collectively, these findings demonstrate that ACX-02 reshapes the balance between facilitating and suppressive pathways, amplifying R_P_-dependent mediation while attenuating vascular constraints. These concurrent sleep-vascular interactions provide a mechanistic basis for enhanced glymphatic clearance under ACX-02 treatment.

A sensitivity analysis of the **Slow Wave** Bayesian model, in which mediator and suppressor effects were re-estimated after sequential removal of individual variables, demonstrated that these pathways operate largely independently, supporting the robustness and modularity of the inferred clearance mechanisms (**Supplementary Figure 4**).

DEX increased plasma Aβ42/Aβ40 and %p-tau217 to a lesser extent than ACX-02, and these changes in the *Dexmedetomidine Trial* were not significant relative to placebo (**Table 2**). Because increases in Aβ42/Aβ40 and %p-tau217 may reflect either enhanced clearance or reduced synaptic-metabolic production¹LL²L, we examined whether the modest DEX-associated shifts were consistent with a production-mediated mechanism (**Supplementary Methods and Results** *Pharmacodynamic mediators and suppressors of DEX treatment effect on AD biomarker clearance to plasma*; **Supplementary Table 5**). Mediation analyses indicated that DEX primarily reduced synaptic-metabolic production, whereas ACX-02 produced a clearance-dominant profile. These findings support a model in which reduced production and enhanced glymphatic clearance represent separable but complementary components of proteostasis modulation that, under optimized vascular conditions with ACX-02, act together to increase net removal of aggregation-prone species from the brain.

### Treatment effect on sleep macrostructure and microstructure

In the *Dexmedetomidine Trial*, DEX treatment increased N2 sleep duration (placebo, 78.24L±L17.38Lmin; treatment, 123.67L±L22.79Lmin; PL=L0.001, linear mixed-effects model) and decreased REM sleep duration (placebo, 42.92L±L15.61Lmin; treatment, 29.28L±L16.33Lmin; PL=L0.004, linear mixed-effects model), with no significant effects on N1, N3, or WASO (**Supplementary Table 2**).

In the *ACX-02 Trial*, treatment similarly increased N2 duration (placebo, 106.4L±L31.2Lmin; treatment, 126.7L±L23.6Lmin; PL=L0.022, linear mixed-effects model) but did not significantly alter REM sleep duration (placebo, 25.0L±L20.3Lmin; treatment, 30.4L±L16.3Lmin; PL=L0.312, linear mixed-effects model). ACX-02 additionally reduced WASO (placebo, 26.3L±L25.5Lmin; treatment, 10.8L±L9.2Lmin; PL=L0.004, linear mixed-effects model), with no effects on N1 or N3 duration (**Supplementary Table 3**).

Both trials exhibited marked increases in slow-wave counts during N2 sleep (DEX: placebo, 69L±L45; treatment, 244L±L190; PL=L0.009, linear mixed-effects model; ACX-02: placebo, 150L±L157; treatment, 479L±L287; PL<L0.001, linear mixed-effects model), without significant changes in N3 sleep. These increases exceeded the proportional expansion of N2 duration, resulting in significantly elevated N2 slow-wave density under both treatments (DEX: placebo, 1.60L±L0.91Lmin⁻¹; treatment, 3.65L±L2.63Lmin⁻¹; PL=L0.009, linear mixed-effects model; ACX-02: placebo, 2.34L±L1.78Lmin⁻¹; treatment, 7.17L±L4.67Lmin⁻¹; PL<L0.001, linear mixed-effects model). Consistent with these observations, slow-wave count emerged as a significant mediator of ACX-02–induced changes in AD biomarker clearance, whereas N2 duration alone did not, indicating as expected that N2 is a poor surrogate for slow-wave activity. Notably, ACX-02 did not significantly affect REM sleep duration or microarchitecture. State occupancy probabilities across wake (W), N1, N2, N3, and REM during the 4-hour sleep period under DEX and the 4-hour 15-minute sleep period under ACX-02 relative to placebo are shown in **Supplementary Figure 6**.

## DISCUSSION

In the present study, we demonstrated that ACX-02, a fixed-dose combination of the centrally acting α2A-adrenergic agonist DEX and a peripherally acting α1-adrenergic agonist midodrine, engaged key physiological determinants of glymphatic function and increased plasma mass-balance indices consistent with enhanced brain-to-blood clearance of Aβ and tau in healthy older adults. In contrast to DEX alone, which increased slow waves, but induced systemic hypotension and did not reduce brain parenchymal resistance to fluid flow, ACX-02 increased EEG slow waves, maintained systemic blood pressure, increased cerebral vasomotor compliance, and reduced brain parenchymal resistance to fluid flow. Bayesian mediation analysis demonstrated that this physiological engagement resulted in measurable changes in plasma Aβ and tau dynamics, indicating that pharmacological enhancement of glymphatic transport can increase the brain-to-blood clearance of Aβ and tau.

Prior experimental studies in animals/rodents have demonstrated that systemic treatment with α2-agonists including xylazine and DEX enhanced glymphatic function^10,21,23^, leading to the suggestion that DEX represents a leading candidate as a clinically relevant enhancer of glymphatic function^37^. Thus, we were surprised to observe that despite its strong impact in increasing slow waves, DEX alone did not reduce brain parenchymal resistance to fluid flow^17^, despite prior studies in rodents suggesting that peripheral α2 agonism increases extracellular volume fraction^10^. Similarly, the absence of an appreciable effect of DEX alone on plasma Aβ and tau dynamics consistent with increased glymphatic clearance^18^, suggested that this drug was not having the anticipated impact on glymphatic clearance.

Analyzing data from the *Dexmedetomidine Trial*, we noted that the degree of systemic hypotension that accompanied DEX infusion may trigger compensatory cerebral autoregulatory vasodilation that could close off perivascular fluid transport pathways, keeping parenchymal resistance to fluid flow too high to permit increased glymphatic clearance of Aβ and tau. The discrepancy in the findings from the DEX trial with prior animal experiments can be explained by the potential for DEX to cause both cerebral and systemic vasoconstriction at doses greater than that used in the DEX trial, that was designed to follow FDA dosing guidance of doses routinely used in the clinical setting^38^. DEX used in animal experiments was typically administered as a single intraperitoneal injection or as a subcutaneous infusion^23^. Although plasma DEX concentrations were not measured in the animal experiments, it was likely that the plasma dose of DEX achieved in experiments exceeded normal clinical levels and reached plasma concentrations that could produce systemic vasoconstriction. In a study performed in rats, peripheral administration of pharmacological vasodilators reduced systemic blood pressure, and blocked CSF tracer influx from the CSF into brain tissue^31^. Within the present study, under DEX alone, systemic hypotension and prolongation of PTT*_cereb_* were consistent with compensatory cerebral autoregulatory vasodilation and increased cerebral blood volume.

These findings provided a critical perspective when evaluating interventions to enhance glymphatic function in clinical populations and demonstrated that enhancing slow waves alone was not sufficient to enhance the glymphatic clearance of Aβ and tau. If the intervention does not engage sleep-associated declines in parenchymal resistance or impairs other determinants of glymphatic transport through peripheral vascular or other off-target effects, its potential impact on glymphatic function may be masked.

We tested the hypothesis that systemic arterial blood pressure was an important modulator of glymphatic function by examining the actions of ACX-02, a fixed-dose combination of DEX iv and oral midodrine. Because midodrine, an α1-agonist, does not cross the blood-brain barrier, we hypothesized that this combination would maintain the effects of central α2-adrenergic agonism on glymphatic transport, while blocking the peripheral vascular effects of DEX. Consistent with the hypothesis, ACX-02 did not evoke the degree of systemic hypotension observed with DEX alone. In the absence of systemic hypotension, ACX-02 engaged three convergent physiologies central to glymphatic transport: (i) it reduced parenchymal resistance, consistent with expansion of the brain’s extracellular compartment^10^; (ii) it increased EEG slow waves, which in animal models is a driver of glymphatic transport^21,39^; and (iii) it altered cerebrovascular pulsatility indexed by PTT*_cereb_*, consistent with preserved or enhanced vasomotor oscillations^32^. With this engagement, ACX-02 increased plasma Aβ42/Aβ40 and %p-tau217 mass ratios by approximately 9%–10% relative to placebo, a pattern that pharmacokinetic analysis suggested was consistent with preferential clearance of aggregation-prone species rather than increased solute release^18^.

Bayesian multivariate mediation analyses indicate that ACX-02 not only activates clearance-related physiological processes but also alters system receptivity to them. Across models designed to mitigate collinearity, R_P_ and PTT*_cereb_* emerged as both mediators and moderators of treatment effect. R_P_ accounted for a substantially larger fraction of total effect under ACX-02 than under placebo, suggesting a synergistic interaction with other ACX-02-evoked effects.

Conversely, the suppressive influence attributed to PTT*_cereb_*under placebo was functionally reversed under ACX-02 administration, underscoring that the physiological significance of prolonged PTT*_cereb_*may depend on the physiological origin of this change. If PTT*_cereb_*is longer due to the absence of cerebrovascular disease and the preservation of cerebrovascular compliance, this may facilitate perivascular propulsion. However, if PTT*_cereb_* lengthens due to the autoregulatory cerebral vasodilation, it may reflect a state with impaired clearance.

Importantly, beyond slow-wave count and its spectral proxy, EEG slow delta power, no additional dominant mediators or suppressors were identified among autonomic, respiratory, or broader EEG measures. Hypnogram stage durations including N2 and N3 did not mediate clearance despite treatment-related changes, reinforcing that sleep stage time is an imperfect surrogate for sleep microstructural features governing glymphatic transport. Slow-wave density increased disproportionately relative to N2 duration, emphasizing the primacy of microstructure over macrostructure. Moreover, ACX-02 did not reduce REM sleep duration.

Because fluid and solute movement depends on the interplay between driving forces and tissue resistance, changes in parenchymal resistance can amplify—or compound—the effects of physiological drivers of glymphatic transport, including neuronal and vasomotor oscillations.

This suggests that sleep may reflect a state in which various physiological factors – neural activity patterns, cerebrovascular dynamics, parenchymal resistance – are aligned to maximize clearance efficiency. These interactions may explain the pathophysiologic relationship between age, cardiovascular disease, and cerebrovascular disease as risk factors for impaired glymphatic function and AD. It would also support the early findings that long term treatment with statins, GLP-1 (glucagon-like peptide-1) receptor antagonists, or SGLT2i (sodium-glucose cotransporter-2 inhibitors) that are effective for improving cardiovascular health have also been observed to decrease the risk of AD and AD-related dementias^40,41^. Obstructive sleep apnea is associated with reduced cerebrovascular compliance, sleep disruption (including reduced slow waves and delta power), and hypoxic/hypercapnic cerebral vasodilation. This may also explain its status as a potent risk factor for dementia, including Alzheimer’s disease^42^. Correspondingly, the most effective interventions for enhancing glymphatic function clinically are expected to be those that appropriately engage each of these factors in a clearance-promoting direction.

Treatment with ACX-02 during a 4-hour and 15-minute sleep period increased brain-to-blood Aβ and tau clearance by ∼9%–10%. If sustained across overnight sleep over longer periods, the magnitude of clearance enhancement observed here may have substantial clinical relevance.

By testing the 4-h and 15-min intervention during the wake period, we also demonstrated that glymphatic clearance is modifiable independent of endogenous circadian rhythms. Cross-sectional biomarker and imaging studies link steady-state shifts in plasma Aβ42/Aβ40 ratios to amyloid PET centiloid (CL) burden and Aβ accumulation kinetics^43^. Extrapolating from these relationships, a persistent ∼10% increase in clearance would reduce the net rate of Aβ accumulation compatible with a multi-year delay in reaching pathological Aβ thresholds, given established accumulation rates of ∼3.0 CL per year^44^. The effectiveness of a single exposure to ACX-02 for increasing glymphatic clearance was demonstrated in subjects without risk factors for AD and without a history of sleep distubances. It is possible that ACX-02 could exert greater effects among individuals with risk factors for AD or sleep pathology or when it is administered in repeated doses over time. Furthermore, the clearance effect of ACX-02 during overnight sleep in an elderly cohort may exceed that observed in this sleep-deprived study, as sleep deprivation would be expected to reduce sleep onset latency and WASO under placebo sleep. The full treatment potential of ACX-02 will require validation in longitudinal studies, among at risk patient populations, and in Aβ- and tau-positive populations.

Targeting glymphatic clearance may be complementary to monoclonal antibody therapies. By enhancing endogenous removal of Aβ and tau, ACX-02 may reduce required antibody exposure, mitigate Amyloid-Related Imaging Abnormality (ARIA) risk in APOE ε4 carriers and address tau species that are not directly targeted by anti-Aβ immunotherapies. Furthermore, investigating the actions of ACX-02 has provided a more nuanced understanding of the physiologic mechanisms governing glymphatic function and may provide or support additional therapeutic targets for intervention. A detailed understanding of pathophysiologic factors that could lead to impaired glymphatic function may also increase the ability to identify and diagnose individuals at risk for AD before the onset of symptoms. For example, the ability to measure and quantify increases in PTT*_cereb_*or decreases in Rp during sleep may add to the diagnostic criteria for AD. Given preclinical evidence implicating glymphatic pathways in tau and α-synuclein pathological progression^11–13,45,46^, this approach may also have broader trans-diagnostic implications. Glymphatic function is impaired in animal models of a wide range of dementia and AD non-genetic risk factors, including aging, cerebrovascular dysfunction, traumatic brain injury, diabetes, hypertension, and chronic sleep disruption^47^. The finding that cerebrovascular responses to arterial pressure play a fundamental role in glymphatic drainage provides a mechanistic link between cardiovascular health and brain health. Together, these results suggest that enhancing glymphatic clearance, as observed with ACX-02, may represent a therapeutic strategy for individuals at risk of neurodegenerative proteinopathies driven by Aβ, tau, and α-synuclein aggregation.

Several limitations warrant consideration. Participants were healthy older adults without established Alzheimer’s pathology or sleep disturbances, and biomarker effects were measured after a single intervention. The cross-over order was fixed with treatment administered first to minimize invasive placebo exposure in case treatment was not tolerated, and although a two-week washout and baseline normalization mitigate carryover and drift, residual period effects cannot be fully excluded. The durability of clearance enhancement with repeated nightly dosing, potential physiological adaptation, interaction with plaque and tangle burden, and heterogeneity in vascular and sleep phenotypes require longitudinal investigation.

In summary, ACX-02 represents a pharmacological modulator of glymphatic-linked clearance with pharmacodynamic target engagement and biomarker-validated endpoints in humans. Our findings support a mechanistic model in which effective glymphatic enhancement requires coordinated engagement of interstitial exchange capacity, sleep microstructure, and vascular pulsatile dynamics in the presence of cerebrovascular autoregulatory states that support perivascular flow. These data establish clearance augmentation as a tractable therapeutic strategy and justify clinical evaluation of sustained glymphatic modulation in neurodegenerative disease populations characterized by impaired proteostasis.

## CODE AVAILABILITY

Code used for the analysis and to produce the figures will be made available on Zenodo.

## Data Availability

All data supporting this study will be made available with Institutional Review Board (IRB) approval and a Data Use Agreement. This ensures compliance with participants’ informed consent and permits non-commercial use for independent validation, publication, and the sharing of new findings.

## ACKNOWLEDGEMENTS

This work was funded by Applied Cognition.

## COMPETING INTERESTS

The authors PD, LG, AC, JJI declare the existence of financial and incentive stock options competing interests. BPL is an advisor to Applied Cognition and declares the existence of incentive stock option competing interests. KY is an advisor to C2N Diagnostics and receives stock options. The remaining authors declare no competing interests.

## SUPPLEMENTARY METHODS AND RESULTS

### Derivation of mass balance ratios

To investigate the treatment versus placebo effect on the clearance of Alzheimer’s disease (AD) biomarker peptides from the interstitial fluid (ISF) into plasma, we focus on the mass cleared to plasma after the intervention relative to before, as this ratio is not influenced by confounding changes in plasma volume during the intervention period. Equation 1 defines this ratio for Aβ42.

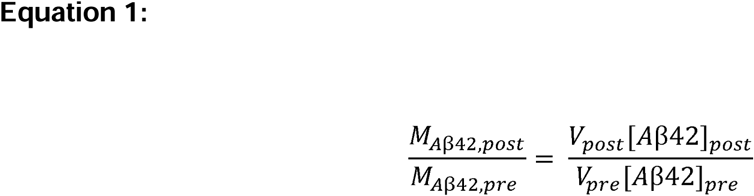

By substituting plasma volume (V_*_) with the expression:

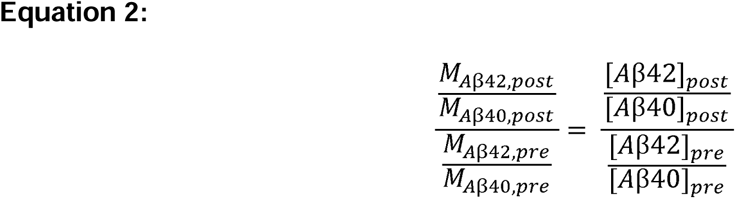

into **Equation 1**, we obtain:

**Equation 2** demonstrates that the ratio of Aβ42/Aβ40 concentrations in plasma post-intervention versus pre-intervention directly reflects the change in total plasma mass of Aβ42 relative to Aβ40. Importantly, this ratio is independent of any changes in plasma volume during the intervention period. The same principle also applies to %p-tau217.

### Pharmacodynamic mediators and suppressors of DEX treatment effect on AD biomarker clearance to plasma

DEX increased plasma Aβ42/Aβ40 and %p-tau217 to a lesser extent than ACX-02 and the changes in the *Dexmedetomidine Trial* were not statistically significant relative to placebo (**Table 2**). Given prior evidence that increases in these plasma ratios may reflect either enhanced clearance or reduced synaptic-metabolic production^18,29^, we sought to determine whether the modest DEX-associated shifts were more consistent with a production-mediated mechanism.

To address this, we constructed Bayesian multivariate linear mixed mediation models paralleling those developed for ACX-02. The joint models evaluated candidate mediators drawn from the same eight physiological categories, including only variables exhibiting effect sizes >0.5 SD under DEX treatment (**Supplemental Table 2**), and excluding variables that were strongly collinear. All mediators and suppressors were standardized (mean 0, SD 1), and treatment was centered (–0.5 placebo, 0.5 DEX). Models included age, sex, and APOE ε4 status as fixed effect covariates, participant ID as a random intercept, and a biomarker-specific random slope to account for the multivariate amyloid and tau endpoints. To address multicollinearity among sleep-related measures, three structurally identical models (**Slow**, **EEG**, **Hypno**) were constructed, each incorporating one sleep-variable set alongside PTT*_cereb_*, LF/HF ratio, respiratory rate (RR), and EtCO₂. Recall that the latter three variables did not meet inclusion criteria in the ACX-02 mediation analysis.

Across all three DEX mediation models, a consistent but non-significant pattern emerged (**Supplementary Figure 5** and **Supplementary Table 5**). Wake after sleep onset (WASO), REM sleep duration, LF/HF ratio, RR, and EtCO₂ appeared as positive mediators of DEX’s effect on plasma Aβ42/Aβ40 and %p-tau217. DEX reduced each of these variables (**Supplemental Table 2**). Reductions in WASO and REM duration are associated with decreased synaptic-metabolic activity; reductions in RR and EtCO₂ reflect decreased systemic metabolic demand; and reductions in LF/HF ratio indicate a shift toward parasympathetic predominance and reduced sympathetic tone. Collectively, these physiological changes are consistent with reduced synaptic-metabolic production of Aβ and tau, supporting the interpretation that DEX’s modest plasma ratio increases were driven primarily by decreased production rather than enhanced glymphatic clearance.

In contrast, variables that mediated clearance under ACX-02, including slow-wave count, EEG slow delta power, and N2 sleep duration, were increased by DEX but functioned as suppressors rather than mediators in these models. This pattern is mechanistically coherent because while increases in slow-wave–related microstructure may promote interstitial transport, DEX-induced hypotension and associated vascular changes likely constrained perivascular CSF influx, thereby limiting clearance engagement. Under these conditions, slow-wave–associated increases in synaptic-metabolic activity may partially offset production-suppressive effects, attenuating net biomarker shifts.

Taken together, mediation analyses under DEX are most consistent with a production-dominant mechanism, whereas ACX-02 produces a clearance-dominant profile. These findings support a model in which reduced synaptic-metabolic production and enhanced glymphatic clearance represent separable yet synergistic effects of proteostasis modulation. Under optimized vascular conditions, as achieved with ACX-02, production reduction and clearance enhancement likely operate together to augment net removal of aggregation-prone species from the brain.

### Plasma AD biomarker assessment

The *APOE* genotyping, Ab and tau plasma biomarkers were analyzed using mass spectrometry by C_2_N Diagnostics^25,26,48^. The sample collection procedure was provided by C_2_N Diagnostics. Blood draw was performed through the IV-line (minimum 22-gauge to minimize red blood cell hemolysis). A total of 10 ml of blood was drawn into a K_2_EDTA Vacutainer. The blood was centrifuged for 15 min using a swinging bucket rotor at 500-700 x g with the brake on.

Immediately after centrifugation, four 1.0 mL plasma samples were aliquoted into four Sarstedt 2.0 ml Micro Tubes without disrupting the plasma/cell interface when transferring plasma. A calibrated air-displacement hand-held pipette with a polypropylene pipette tip was used. After aliquoting plasma into the Sarstedt Micro Tubes, the tubes were immediately capped and immediately frozen at −80°C. When the tubes were ready to be shipped to C_2_N Diagnostics, they were packed into a plastic zip-lock bag with plenty of dry ice, placed in an absorbent towel and cryobox, and express couriered to C_2_N Diagnostics priority overnight.

### Signal Processing – Electroencephalography

The ADS1299 uses analog-to-digital converters (ADCs) sampling at 1 Msps, with digital filtering (third-order sync filter) and decimation resulting in an output rate of 250 sps. Anti-aliasing was implemented with first-order differential low-pass filters at 16.5 kHz (−3dB frequency), and first-order single-ended low-pass filters at 330 kHz for further attenuation of RF interference. The raw EEG tracings from the device were notch-filtered at 60 Hz using a second-order infinite impulse response (IIR) digital filter (Python scipy.signal.iirnotch). This filter was applied to the signal in both forward and backward directions using zero-phase filtering (Python scipy.signal.filtfilt). The signal was then bandpass filtered between 0.3 Hz and 50 Hz using a finite impulse response (FIR) filter with a Hann window (Python scipy.signal.firwin). The low-pass filter length was 501, and the high-pass filter length was 2,401.

During the observation periods, participants were not allowed to touch devices connected to power outlets. Non-physiological sources of artifact included excessive head motion, which introduced signal noise at the electrode–skin interface. Physiological artifacts included electrooculogram (EOG), electrocardiogram (ECG), and electromyogram (EMG) signals. EEG signals were partitioned into time-aligned 30-second epochs. Epochs were removed if they contained a peak-to-peak signal amplitude exceeding 350 µV or maximum power in the Welch power spectrum exceeding 1,000 µV²/Hz. Additionally, IMU signals from the device’s left and right in-ear sensors were used to filter for excessive motion by excluding epochs in which peak-to-peak acceleration on any x-, y-, or z-axis exceeded 100 mG during sleep or 200 mG during wakefulness.

Hypnogram staging for each 30-second epoch was performed using automated sleep scoring (Python yasa 0.6.3 SleepStaging), trained and validated on 3,000 nights of data from the National Sleep Research Resource^49^. Because the algorithm uses a single EEG derivative, the device’s single in-ear transcranial derivative was used for hypnogram staging.

Power spectral density (PSD) for each 30-second epoch was computed using Welch’s method^50^ (Python scipy.signal.welch) with 10-second segments and 50% overlap. Relative power was calculated for standard frequency bands—slow delta (0.5 – 1 Hz), delta (1–4 Hz), theta (4–8 Hz), alpha (8–12 Hz), sigma (12–15 Hz), beta (15–30 Hz), and low gamma (30–50 Hz)—normalized to the total power in the power spectrum. Simpson’s rule was applied to compute total and band-specific powers from the PSD. EEG power during sleep was separately averaged across REM epochs and across NREM epochs (N2 and N3).

## SUPPLEMENTARY TABLES AND FIGURES

**Supplementary Figure 1:**
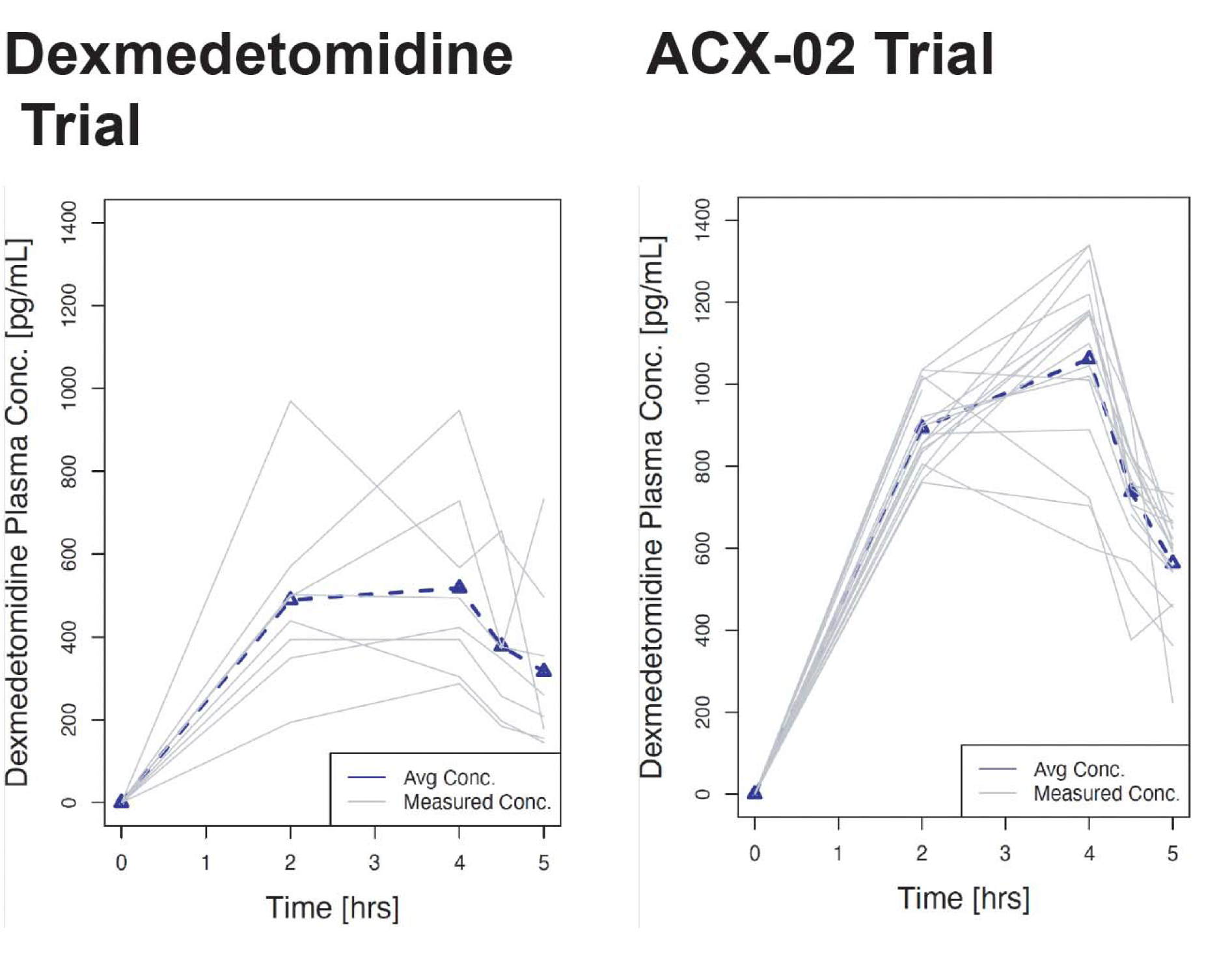
Plasma concentrations of DEX during the treatment study visit. Plasma DEX concentrations measured at infusion onset, 2 h after infusion onset, at infusion termination and at 30 min and 1 h post-infusion for the *Dexmedetomidine Trial* and *ACX-02 Trial*. In the *Dexmedetomidine Trial*, infusion was initiated at 0.6 μg/kg/h and titrated to avoid apnea and to maintain mean arterial pressure ≥60 mmHg, heart rate ≥45 bpm, and SpO₂ ≥92%. In the *ACX-02 Trial*, DEX was infused at a fixed rate of 0.7 μg/kg/h throughout the 4-h 15-min treatment period.

**Supplementary Figure 2:**
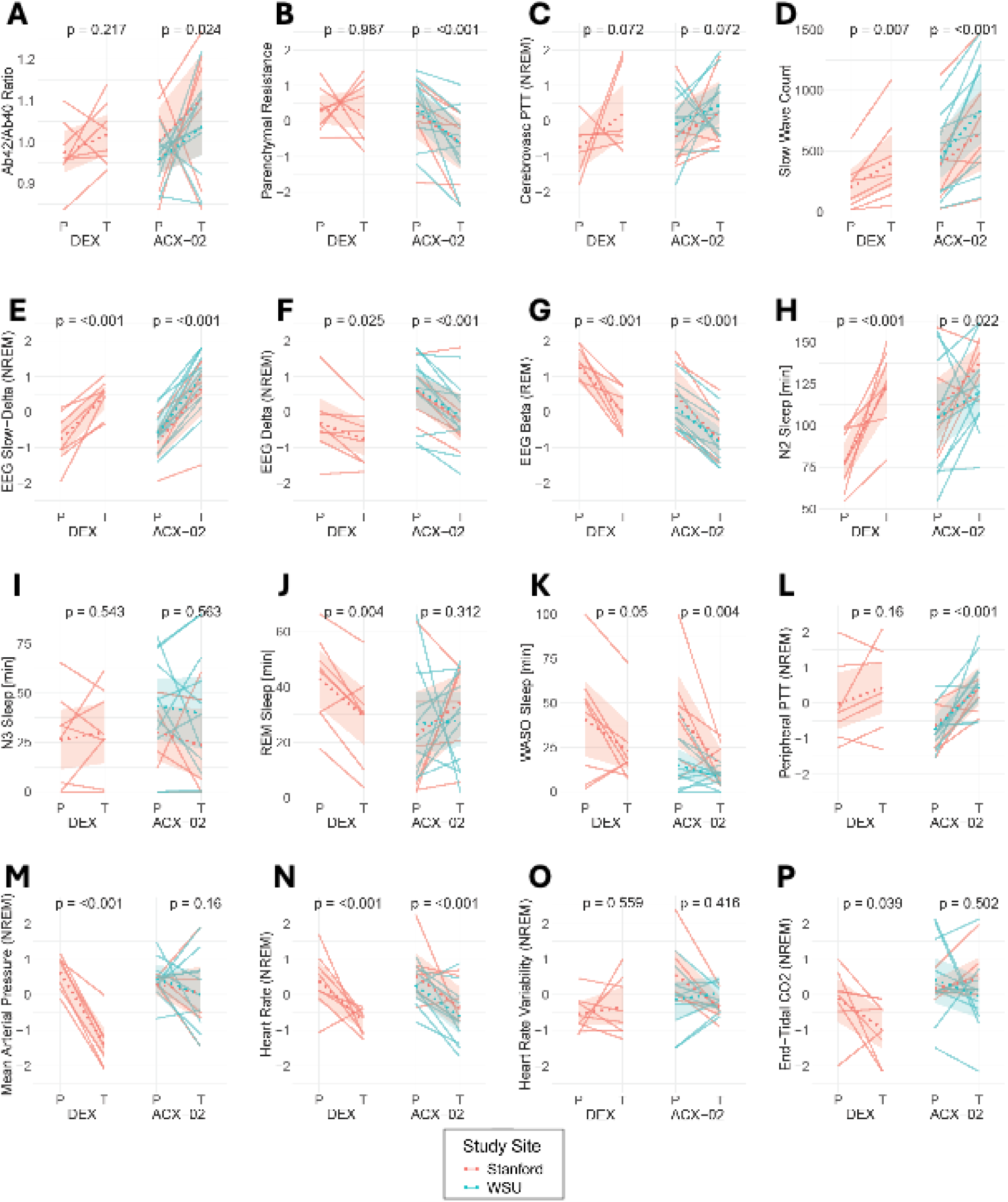
Effects of DEX and ACX-02 on sleep-linked glymphatic features and on plasma biomarkers of Alzheimer’s disease. Comparisons between placebo and treatment pharmacodynamic and biomarker values, averaged over the sleep opportunity and normalized to the corresponding baseline, are shown for both the *Dexmedetomidine* and *ACX-02 Trials* for **(A)** Aβ42/Aβ40 ratio, **(B)** Parenchymal Resistance, **(C)** Cerebrovascular PTT (NREM), **(D)** Slow Wave Count, **(E)** EEG Slow-Delta Power (NREM), **(F)** EEG Delta Power (NREM), **(G)** EEG Beta Power (REM), **(H)** N2 Sleep Duration, **(I)** N3 Sleep Duration, **(J)** REM Sleep Duration, **(K)** WASO Duration, **(L)** Peripheral PTT (NREM), **(M)** Mean Arterial Pressure (NREM), **(N)** Heart Rate (NREM), **(O)** Heart Rate Variability (NREM), **(P)** End-Tidal CO_2_ (NREM). No notable differences were observed between study sites in the *ACX-02 Trial*. Average site values are indicated by dotted lines. Reported two-sided p-values for fixed effects were obtained using Wald t-tests with Satterthwaite degrees of freedom from a linear mixed-effects model. Significance tests were not adjusted for multiple comparisons. Placebo (P) and Treatment (T).

**Supplementary Figure 3:**
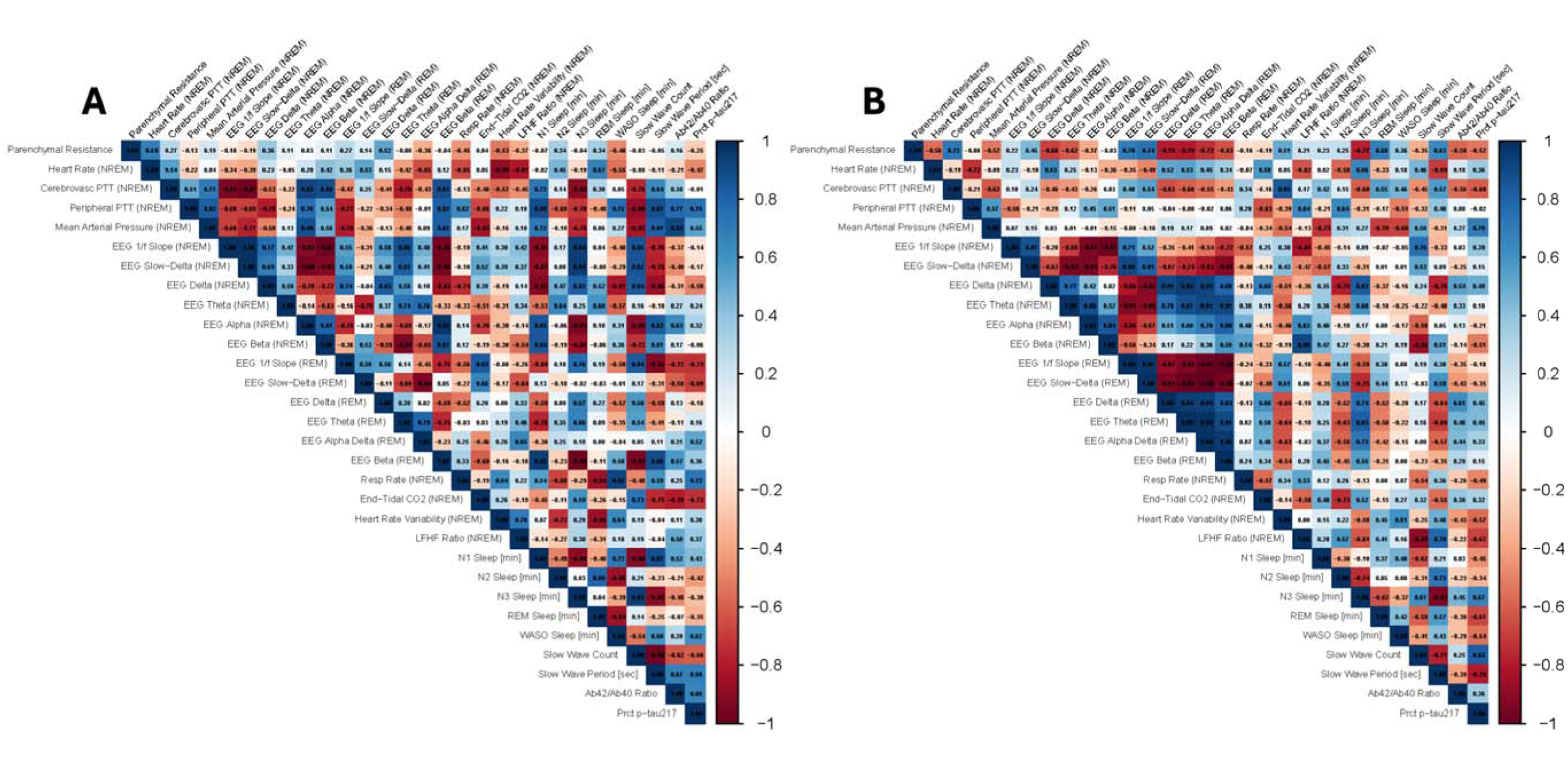
Correlations of pharmacodynamic and AD biomarker values during DEX and ACX-02 treatment. The between-patient correlations of physiological measurements during **(A)** DEX treatment and **(B)** ACX-02 treatment are shown. The key drivers of glymphatic clearance, R_P_, PTT*_cereb_*, slow-waves, and EEG spectral bands show distinct differences between DEX and ACX-02 with ACX-02 inducing stronger correlations among EEG spectral bands and R_P_ and weaker correlations with PTT*_cereb_*.

**Supplementary Figure 4:**
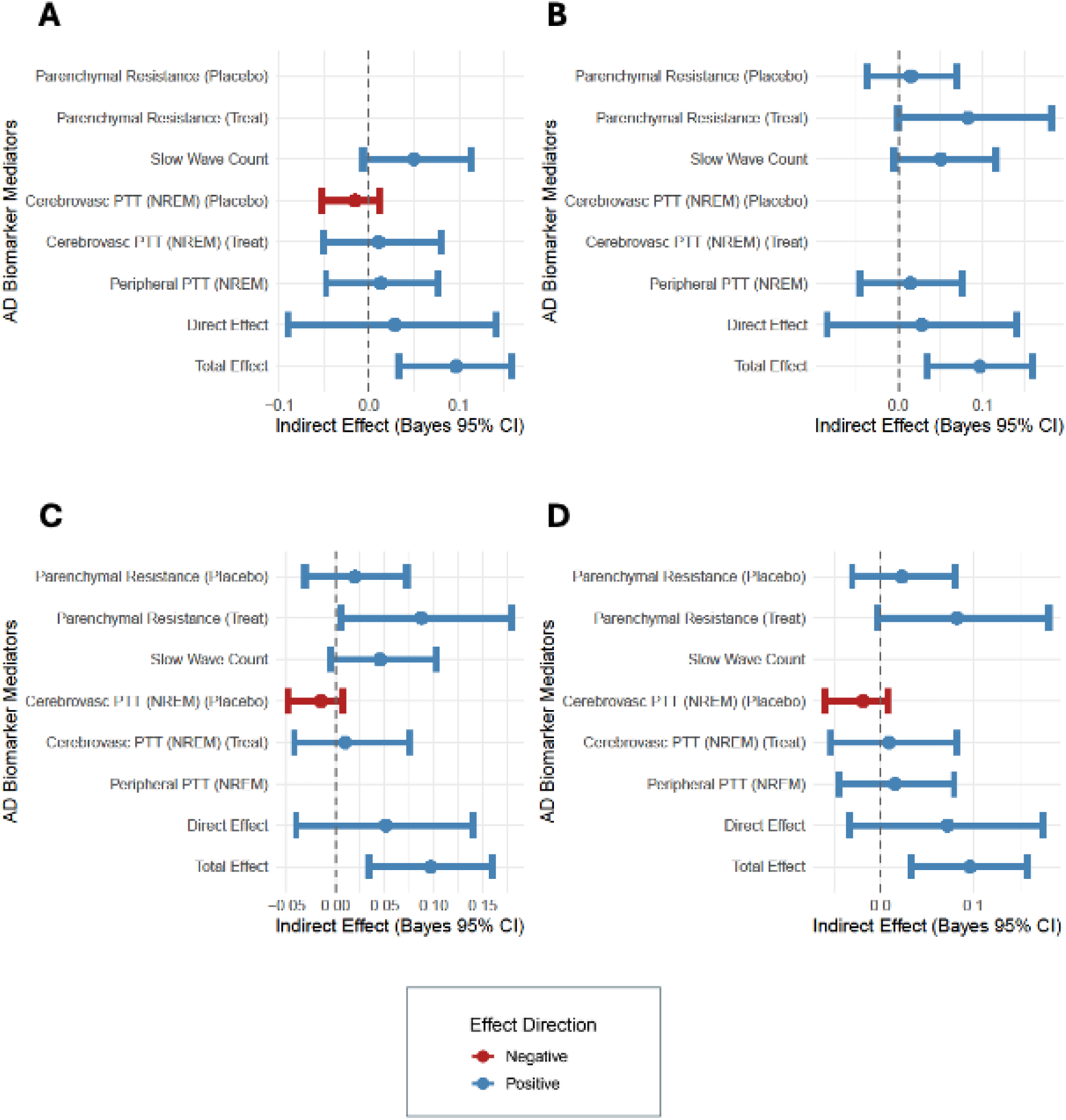
Sensitivity analysis for indirect mediation effect, direct effect and total effect of ACX-02 on glymphatic clearance. Sensitivity analysis of the Bayesian linear mixed model involved sequentially dropping **(A)** Parenchymal Resistance, **(B)** Cerebrovascular PTT (NREM), **(C)** Peripheral PTT (NREM) and **(D)** Slow Wave Count and re-estimating the model. All selected variables were standardized (mean 0, SD 1) prior to Bayesian mediation modeling using weakly informative priors. A total of 8,000 posterior simulations were used to estimate mean effects and 95% Bayesian confidence intervals (Bayes 95% CI).

**Supplementary Figure 5:**
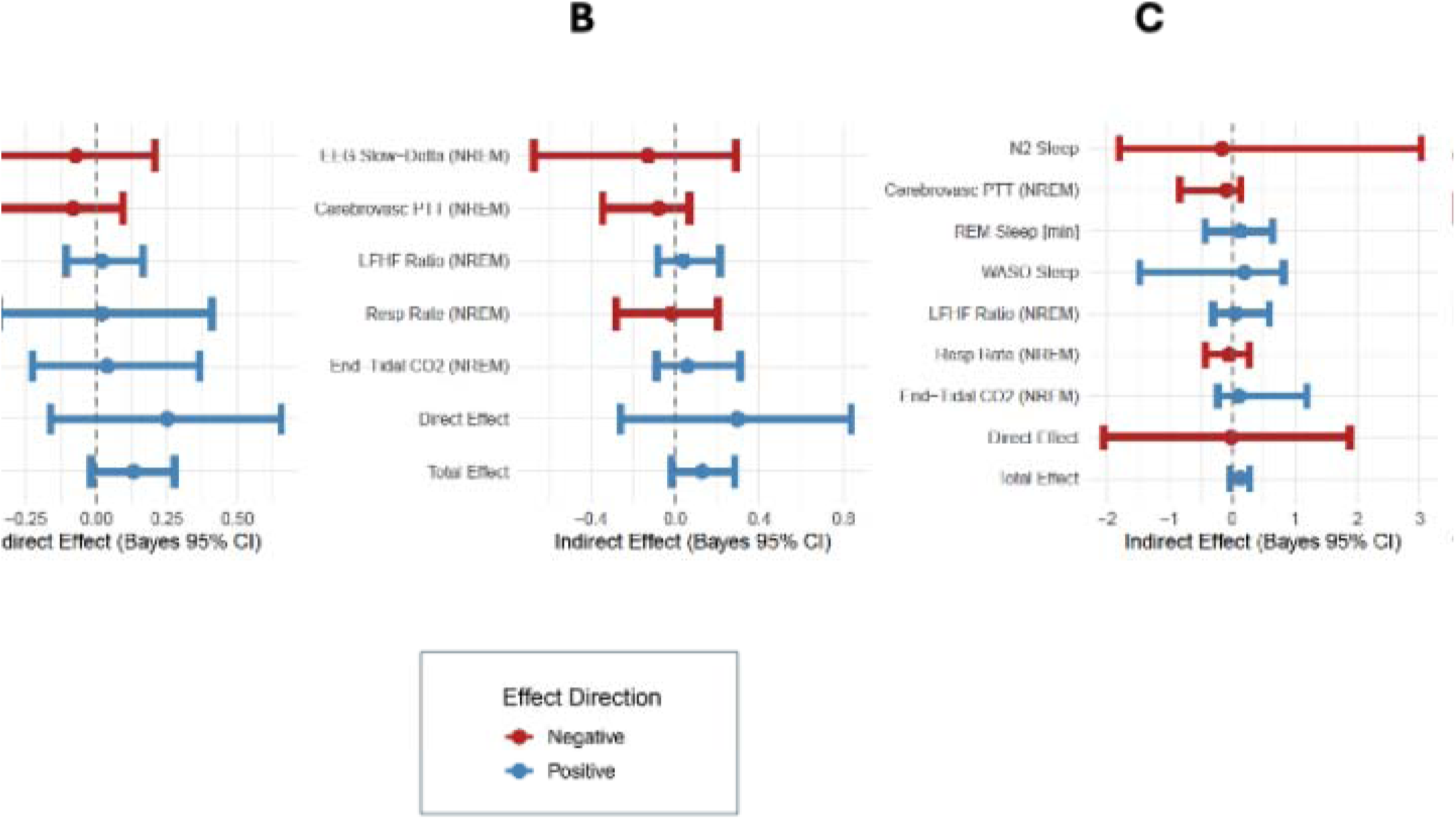
Indirect mediation effect, direct effect and total effect of DEX on glymphatic clearance in the Dexmedetomidine Trial. Several physiological variables continuously monitored during the DEX intervention exhibited significant treatment responses (standardized effect size >0.5 SD) and were therefore evaluated as candidate mediators or suppressors of DEX’s effect on plasma Aβ42/Aβ40 and %p-tau217. These included: (i) cerebrovascular pulsatile dynamics (cerebrovascular PTT during NREM); (ii) slow-wave microstructure (slow-wave count); (iii) EEG spectral power (slow delta power during NREM); (iv) hypnogram stage durations (N2, REM, and WASO); (v) autonomic balance (LF/HF ratio during NREM); and (vi) respiratory physiology (respiratory rate and end-tidal CO₂ during NREM). All selected variables were standardized (mean 0, SD 1) prior to Bayesian mediation modeling using weakly informative priors. Because slow-wave count, EEG power bands, and hypnogram durations were strongly intercorrelated, three separate Bayesian multivariate linear mixed models were constructed to mitigate collinearity. Each model included cerebrovascular PTT (NREM), LF/HF ratio (NREM), respiratory rate (NREM), and EtCO₂ (NREM), and differed only in the inclusion of one sleep-related domain: **(A)** slow-wave count, **(B)** NREM EEG slow delta power, or **(C)** hypnogram stage durations. Models incorporated random intercepts for participant and biomarker-specific random slopes. A total of 8,000 posterior simulations were used to estimate mean effects and 95% Bayesian confidence intervals (Bayes 95% CI). Because DEX increased plasma Aβ42/Aβ40 and %p-tau217, mediation directionality was defined relative to whether treatment-induced changes in a physiological variable predicted concordant versus opposing shifts in plasma ratios. LF/HF ratio, respiratory rate, EtCO₂, WASO, and REM duration were all reduced by DEX. These variables emerged as positive mediators, indicating that their reduction was associated with increased plasma ratios. Physiologically, decreases in these measures are consistent with reduced systemic and synaptic-metabolic activity, supporting the interpretation that DEX increased plasma biomarker ratios primarily through reduced production of amyloid and tau^18^. By contrast, DEX increased cerebrovascular PTT, slow-wave count, NREM EEG slow delta power, and N2 duration. These variables functioned as suppressors in mediation models, suggesting that their treatment-induced increases opposed the net plasma ratio effect. This pattern implies that although DEX enhanced slow-wave–related microstructure, concurrent vascular and cerebrovascular changes limited effective glymphatic engagement, shifting the net effect toward production reduction rather than clearance enhancement. This contrasts with ACX-02, in which similar sleep-related increases acted as mediators within a vascular state permissive to clearance. Together, these analyses support a production-dominant mechanism for DEX, distinct from the clearance-dominant profile observed under ACX-02.

**Supplementary Figure 6:**
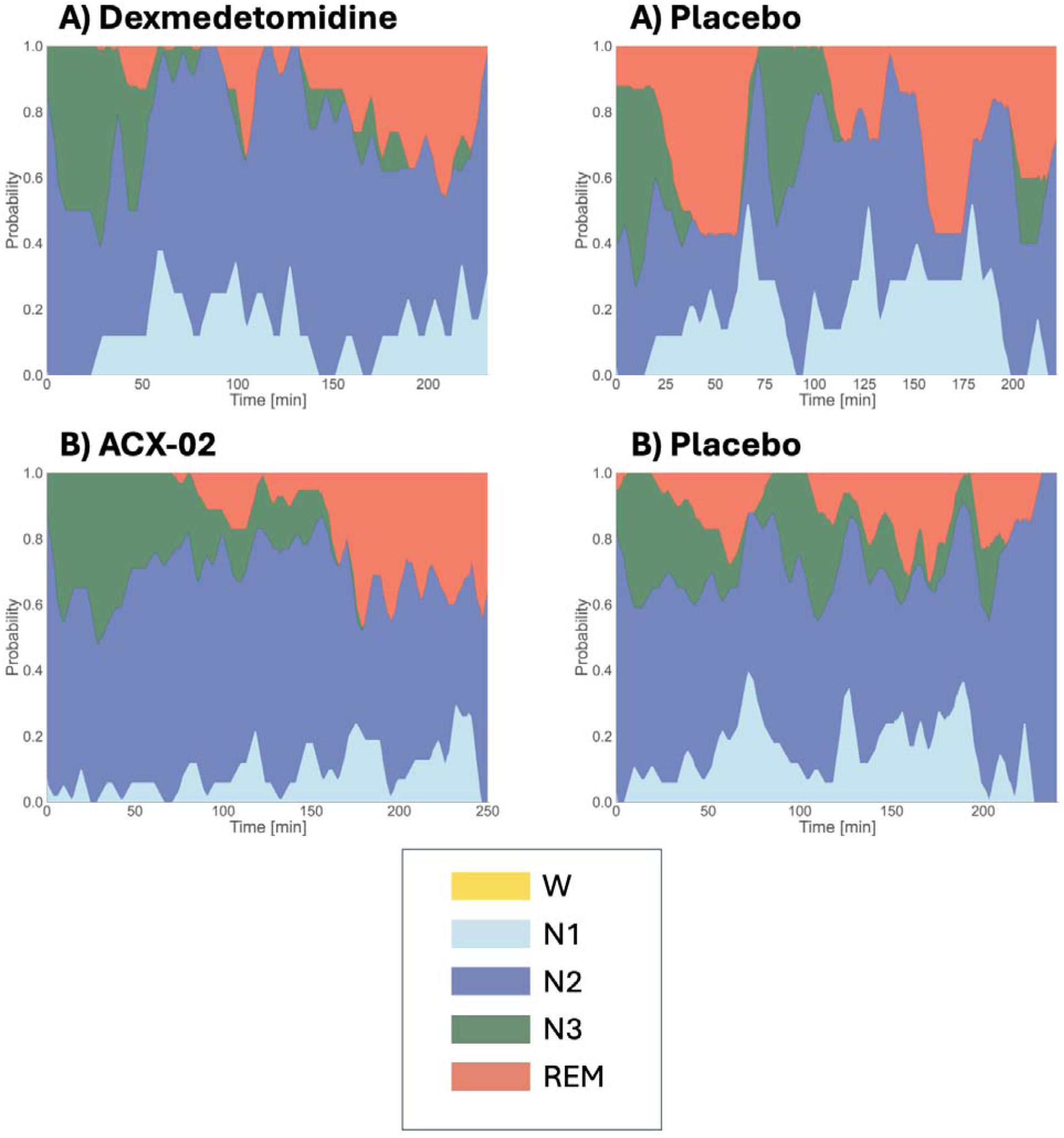
Sleep macrostructure during DEX and ACX-02 treatment versus placebo. The probability per epoch of participants being in one of the five sleep stages, wake (W), N1, N2, N3, and rapid eye movement (REM), is shown for the sleep period under **(A)** DEX and **(B)** ACX-02 treatment compared to their respective placebo.

---

**Supplementary Table 1:**
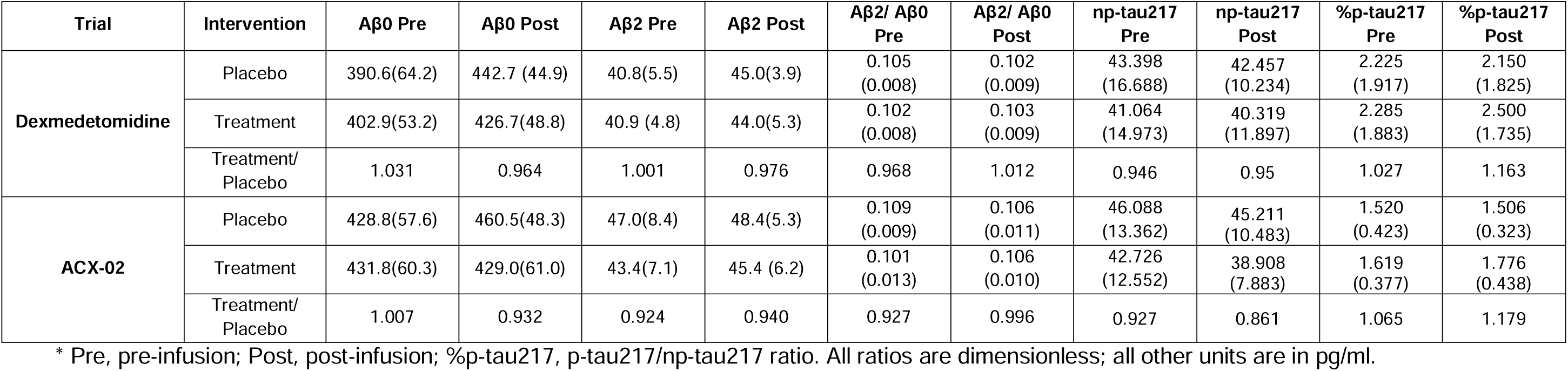
Plasma AD biomarker levels (mean, SD) and ratios pre and post placebo and treatment infusion for the *Dexmedetomidine* and *ACX-02 Trials*\*.

| Trial | Intervention | Aβ0 Pre | Aβ0 Post | Aβ2 Pre | Aβ2 Post | Aβ2/ Aβ0 Pre | Aβ2/ Aβ0 Post | np-tau217 Pre | np-tau217 Post | %p-tau217 Pre | %p-tau217 Post |
| --- | --- | --- | --- | --- | --- | --- | --- | --- | --- | --- | --- |
| **Dexmedetomidine** | Placebo | 390.6(64.2) | 442.7 (44.9) | 40.8(5.5) | 45.0(3.9) | 0.105<br>(0.008) | 0.102<br>(0.009) | 43.398<br>(16.688) | 42.457<br>(10.234) | 2.225<br>(1.917) | 2.150<br>(1.825) |
|  | Treatment | 402.9(53.2) | 426.7(48.8) | 40.9 (4.8) | 44.0(5.3) | 0.102<br>(0.008) | 0.103<br>(0.009) | 41.064<br>(14.973) | 40.319<br>(11.897) | 2.285<br>(1.883) | 2.500<br>(1.735) |
|  | Treatment/<br>Placebo | 1.031 | 0.964 | 1.001 | 0.976 | 0.968 | 1.012 | 0.946 | 0.95 | 1.027 | 1.163 |
| **ACX-02** | Placebo | 428.8(57.6) | 460.5(48.3) | 47.0(8.4) | 48.4(5.3) | 0.109<br>(0.009) | 0.106<br>(0.011) | 46.088<br>(13.362) | 45.211<br>(10.483) | 1.520<br>(0.423) | 1.506<br>(0.323) |
|  | Treatment | 431.8(60.3) | 429.0(61.0) | 43.4(7.1) | 45.4 (6.2) | 0.101<br>(0.013) | 0.106<br>(0.010) | 42.726<br>(12.552) | 38.908<br>(7.883) | 1.619<br>(0.377) | 1.776<br>(0.438) |
|  | Treatment/<br>Placebo | 1.007 | 0.932 | 0.924 | 0.940 | 0.927 | 0.996 | 0.927 | 0.861 | 1.065 | 1.179 |

\* Pre, pre-infusion; Post, post-infusion; %p-tau217, p-tau217/np-tau217 ratio. All ratios are dimensionless; all other units are in pg/ml.

**Supplementary Table 2:** Effect of DEX treatment in the *Dexmedetomidine Trial* on pharmacodynamic measures*.

| Physiological Parameters | Pharmacodynamic Response | Treatment Mean (SD) | Placebo Mean (SD) | Effect Size Estimate | DF | CI | P |
| --- | --- | --- | --- | --- | --- | --- | --- |
| **Sleep-associated modulation of extracellular volume** | Parenchymal Resistance (R<sub>P</sub>) | -0.004 (1.199) | 0.004 (0.839) | -0.007 | 16 | -0.95 – 0.93 | 0.987 |
| **Cerebral vasomotor oscillations** | Cerebrovascular PTT (NREM) | 0.381 (1.031) | -0.509 (0.755) | 0.886 | 14 | -0.09 – 1.86 | 0.072 |
| **Peripheral vascular tone** | Heart Rate (NREM) | -0.669 (0.420) | 0.669 (0.971) | -1.338 | 16 | -2.04 – -0.63 | **0.001** |
|  | Peripheral PTT (NREM) | 0.192 (1.000) | -0.220 (1.031) | 0.432 | 8 | -0.21 – 1.07 | 0.160 |
|  | Mean Arterial Pressure (NREM) | -0.796 (0.551) | 0.910 (0.410) | -1.676 | 8 | -2.02 – -1.33 | **<0.001** |
| **EEG power bands** | EEG 1/f Slope (NREM) | 0.502 (0.916) | -0.502 (0.853) | 1.003 | 8 | 0.54 – 1.47 | **0.001** |
|  | EEG Slow Delta Power (NREM) | 0.702 (0.578) | -0.702 (0.826) | 1.404 | 8 | 0.86 – 1.94 | **<0.001** |
|  | EEG Delta Power (NREM) | -0.266 (0.849) | 0.266 (1.123) | -0.532 | 8 | -0.98 – -0.08 | **0.025** |
|  | EEG Theta Power (NREM) | -0.809 (0.444) | 0.809 (0.671) | -1.618 | 8 | -1.99 – -1.24 | **<0.001** |
|  | EEG Alpha Power (NREM) | -0.812 (0.438) | 0.812 (0.667) | -1.624 | 8 | -2.01 – -1.24 | **<0.001** |
|  | EEG Beta Power (NREM) | -0.303 (0.888) | 0.303 (1.070) | -0.606 | 8 | -1.15 – -0.06 | **0.034** |
|  | EEG 1/f Slope (REM) | 0.813 (0.742) | -0.711 (0.537) | 1.540 | 15 | 1.04 – 2.04 | **<0.001** |
|  | EEG Slow Delta Power (REM) | 0.745 (0.948) | -0.652 (0.435) | 1.447 | 15 | 0.76 – 2.13 | **<0.001** |
|  | EEG Delta Power (REM) | 0.515 (0.775) | -0.450 (0.994) | 0.892 | 7 | 0.20 – 1.59 | **0.019** |
|  | EEG Theta Power (REM) | 0.003 (1.116) | -0.003 (0.965) | 0.209 | 6 | -0.47 – 0.89 | 0.483 |
|  | EEG Alpha Delta Power (REM) | -0.485 (1.107) | 0.425 (0.713) | -0.973 | 7 | -1.73 – -0.22 | **0.018** |
|  | EEG Beta Power (REM) | -0.739 (0.685) | 0.647 (0.757) | -1.412 | 15 | -2.11 – -0.72 | **0.001** |
| **Respiratory measures** | Respiratory Rate (NREM) | -0.457 (1.177) | 0.457 (0.530) | -0.914 | 16 | -1.65 – -0.18 | **0.018** |
|  | End-Tidal CO<sub>2</sub> (NREM) | -0.449 (0.891) | 0.449 (0.942) | -0.899 | 16 | -1.75 – -0.05 | **0.039** |
| **Autonomic nervous system measures** | Heart Rate Variability (NREM) | 0.140 (1.217) | -0.140 (0.785) | 0.281 | 16 | -0.71 – 1.28 | 0.559 |
|  | LFHF Ratio (NREM) | -0.414 (0.601) | 0.414 (1.179) | -0.827 | 16 | -1.66 – 0.00 | **0.050** |
| **Hypnogram sleep stage durations** | N1 Sleep [min] | 23.1 (14.6) | 22.8 (5.4) | 0.350 | 16 | -6.95 – 7.65 | 0.920 |
|  | N2 Sleep [min] | 123.7 (22.8) | 78.2 (17.4) | 45.4 | 8 | 25.24 – 65.6 | **0.001** |
|  | N3 Sleep [min] | 30.2 (22.3) | 26.3 (23.5) | 3.97 | 8 | -10.43 – 18.4 | 0.543 |
|  | REM Sleep [min] | 29.3 (16.3) | 42.9 (15.6) | -13.6 | 8 | -21.6 – -5.69 | **0.004** |
|  | WASO Sleep [min] | 23.1 (21.0) | 40.7 (32.9) | -17.6 | 8 | -35.2 – -0.01 | **0.050** |
| **Slow waves** | Slow Wave Count | 406.6 (338.1) | 202.1 (196.8) | 204.5 | 8 | 75.1 – 334.0 | **0.007** |
|  | Slow Wave Period [sec] | 1.455 (0.074) | 1.354 (0.088) | 0.091 | 8 | 0.05 – 0.13 | **0.001** |
|  | SW Count (N2) [sec] | 243.6 (189.9) | 68.7 (44.8) | 174.9 | 8 | 56.3 – 293.5 | **0.009** |
|  | SW Count (N3) [sec] | 163.0 (162.1) | 133.1 (162.7) | 29.9 | 8 | -42.4 – 102.2 | 0.368 |
|  | SW Density (N2) [cnt/min] | 3.652 (2.634) | 1.596 (0.906) | 2.056 | 8 | 0.663 – 3.448 | **0.009** |
|  | SW Density (N3) [cnt/min] | 8.692 (3.776) | 7.659 (4.648) | 1.620 | 6 | -0.272 – 3.513 | 0.082 |

\* DF, Satterthwaite degrees of freedom; CI, 95% confidence interval; P, linear mixed effects model two-sided *p*-value calculated using a *t*-distribution with Satterthwaite degrees of freedom. Significance tests were not adjusted for multiple comparisons. Response variables were standardized to zero mean and unit variance, with Mean, Effect Size and CI's shown in units of SD except for hypnogram values, slow wave count and slow wave period. **Bolded P values reflect significant associations at P < 0.05**

**Supplementary Table 3:** Effect of ACX-02 treatment on pharmacodynamic measures*.

| Physiological Parameters | Pharmacodynamic Response | Treatment Mean (SD) | Placebo Mean (SD) | Effect Size Estimate | DF | CI | P |
| --- | --- | --- | --- | --- | --- | --- | --- |
| **Sleep-associated modulation of extracellular volume** | Parenchymal Resistance (R<sub>p</sub>) | -0.453 (0.963) | 0.453 (0.835) | -0.902 | 19 | -1.34 – -0.47 | **<0.001** |
| **Cerebral vasomotor oscillations** | Cerebrovascular PTT (NREM) | 0.287 (0.991) | -0.304 (0.943) | 0.572 | 35 | -0.05 – 1.20 | 0.072 |
| **Peripheral vascular tone** | Heart Rate (NREM) | -0.387 (1.030) | 0.387 (0.825) | -0.785 | 17 | -1.20 – -0.37 | **0.001** |
|  | Peripheral PTT (NREM) | 0.568 (0.879) | -0.639 (0.708) | 1.202 | 34 | 0.66 – 1.75 | **<0.001** |
|  | Mean Arterial Pressure (NREM) | -0.204 (1.243) | 0.229 (0.587) | -0.444 | 34 | -1.07 – 0.18 | 0.160 |
| **EEG power bands** | EEG 1/f Slope (NREM) | 0.571 (0.940) | -0.571 (0.696) | 1.189 | 18 | 0.92 – 1.46 | **<0.001** |
|  | EEG Slow Delta Power (NREM) | 0.725 (0.786) | -0.725 (0.572) | 1.440 | 18 | 1.18 – 1.70 | **<0.001** |
|  | EEG Delta Power (NREM) | -0.410 (0.928) | 0.410 (0.918) | -0.790 | 18 | -1.08 – -0.50 | **<0.001** |
|  | EEG Theta Power (NREM) | -0.759 (0.555) | 0.759 (0.729) | -1.476 | 17 | -1.81 – -1.14 | **<0.001** |
|  | EEG Alpha Power (NREM) | -0.773 (0.530) | 0.773 (0.715) | -1.570 | 18 | -1.85 – -1.29 | **<0.001** |
|  | EEG Beta Power (NREM) | -0.386 (0.908) | 0.386 (0.959) | -0.835 | 18 | -1.08 – -0.59 | **<0.001** |
|  | EEG 1/f Slope (REM) | 0.642 (0.992) | -0.682 (0.349) | 1.373 | 18 | 0.93 – 1.82 | **<0.001** |
|  | EEG Slow Delta Power (REM) | 0.675 (0.953) | -0.717 (0.315) | 1.423 | 18 | 0.95 – 1.90 | **<0.001** |
|  | EEG Delta Power (REM) | -0.230 (0.964) | 0.245 (1.009) | -0.557 | 33 | -1.22 – 0.11 | 0.098 |
|  | EEG Theta Power (REM) | -0.505 (1.014) | 0.537 (0.663) | -1.059 | 33 | -1.63 – -0.49 | **0.001** |
|  | EEG Alpha Delta Power (REM) | -0.679 (0.805) | 0.722 (0.602) | -1.408 | 18 | -1.90 – -0.91 | **<0.001** |
|  | EEG Beta Power (REM) | -0.580 (0.777) | 0.616 (0.838) | -1.193 | 15 | -1.44 – -0.95 | **<0.001** |
| **Respiratory measures** | Respiratory Rate (NREM) | -0.151 (1.021) | 0.160 (0.982) | -0.351 | 35 | -0.89 – 0.19 | 0.196 |
|  | End-Tidal CO<sub>2</sub> (NREM) | -0.098 (1.036) | 0.103 (0.981) | -0.202 | 19 | -0.82 – 0.42 | 0.502 |
| **Autonomic nervous system measures** | Heart Rate Variability (NREM) | -0.107 (0.939) | 0.107 (1.074) | -0.213 | 19 | -0.75 – 0.32 | 0.416 |
|  | LFHF Ratio (NREM) | 0.178 (1.247) | -0.188 (0.632) | 0.332 | 16 | -0.28 – 0.95 | 0.268 |
| **Hypnogram sleep stage durations** | N1 Sleep [min] | 21.2 (13.6) | 19.5 (13.9) | 1.038 | 36 | -6.35 – 8.43 | 0.777 |
|  | N2 Sleep [min] | 126.7 (23.6) | 106.4 (31.2) | 19.505 | 19 | 3.10 – 35.91 | **0.022** |
|  | N3 Sleep [min] | 33.5 (30.951) | 38.4 (25.2) | -3.574 | 19 | -16.2 – 9.11 | 0.563 |
|  | REM Sleep [min] | 30.399 (16.3) | 25.0 (20.3) | 5.837 | 36 | -5.70 – 17.37 | 0.312 |
|  | WASO Sleep [min] | 10.8 (9.2) | 26.3 (25.5) | -15.583 | 36 | -25.75 – -5.41 | **0.004** |
| **Slow waves** | SW Count | 764.9 (467.8) | 427.3 (328.8) | 359.2 | 18 | 226.4 – 492.0 | **<0.001** |
|  | SW Period [sec] | 1.390 (0.082) | 1.307 (0.075) | 0.073 | 17 | 0.05 – 0.09 | **<0.001** |
|  | SW Count (N2) [sec] | 478.9 (297.0) | 150.0 (157.4) | 334.9 | 19 | 202.8 – 467.2 | **<0.001** |
|  | SW Count (N3) [sec] | 296.8 (313.4) | 277.3 (241.3) | 34.32 | 18 | -47.5 – 116.1 | 0.390 |
|  | SW Density (N2) [cnt/min] | 7.169 (4.669) | 2.337 (1.782) | 4.956 | 19 | 2.93 – 6.99 | **<0.001** |
|  | SW Density (N3) [cnt/min] | 13.314 (7.866) | 12.668 (4.709) | 2.161 | 14 | -0.73 – 5.05 | 0.131 |

\* DF, Satterthwaite degrees of freedom; CI, 95% confidence interval; P, linear mixed effects model two-sided *p*-value calculated using a *t*-distribution with Satterthwaite degrees of freedom. Significance tests were not adjusted for multiple comparisons. Response variables were standardized to zero mean and unit variance, with Mean, Effect Size and CI's shown in units of SDs except for hypnogram values, slow wave count and slow wave period. **Bolded P values reflect significant associations at P < 0.05**

**Supplementary Table 4:** Pharmacodynamic mediators and suppressors of glymphatic clearance engaged by ACX-02*.

| **Sleep Mediators in Model** | **Mediator or Suppressor** | **Effect Size** | **Fraction of Total Effect (%)** | **95% CI** | **Bayes 1-Sided P value** |
| --- | --- | --- | --- | --- | --- |
| **Slow Wave Count** | Parenchymal Resistance (*Placebo*) | 0.015 | 12.2% | -0.040 – 0.070 | 0.286 |
|  | Parenchymal Resistance (*Treatment*) | 0.081 | 68.1% | -0.004 – 0.185 | **0.031** |
|  | Cerebrovascular PTT (NREM) (*Placebo*) | -0.015 | -13.1% | -0.053 – 0.012 | 0.115 |
|  | Cerebrovascular PTT (NREM) (*Treatment*) | 0.011 | 8.7% | -0.050 – 0.083 | 0.347 |
|  | Slow Wave Count | 0.050 | 42.0% | -0.005 – 0.116 | **0.039** |
|  | Peripheral PTT (NREM) | 0.014 | 11.3% | -0.045 – 0.079 | 0.317 |
|  | **Direct Effect** | 0.030 | 24.9% | -0.082 – 0.141 | 0.298 |
|  | **Total Effect** | 0.119 |  | 0.052 – 0.184 | **<0.001** |
| **EEG Powerband** | Parenchymal Resistance (*Placebo*) | 0.037 | 31.3% | -0.049 – 0.130 | 0.186 |
|  | Parenchymal Resistance (*Treatment*) | 0.097 | 81.0% | -0.023 – 0.231 | 0.056 |
|  | Cerebrovascular PTT (NREM) (*Placebo*) | -0.028 | -23.8% | -0.085 – 0.009 | 0.064 |
|  | Cerebrovascular PTT (NREM) (*Treatment*) | 0.013 | 10.9% | -0.064 – 0.104 | 0.350 |
|  | EEG Slow Delta Power (NREM) | 0.156 | 130.8% | -0.068 – 0.390 | 0.084 |
|  | EEG Delta Power (NREM) | -0.039 | -32.7% | -0.139 – 0.057 | 0.205 |
|  | EEG Beta Power (REM) | -0.037 | -30.6% | -0.145 – 0.067 | 0.235 |
|  | Peripheral PTT (NREM) | 0.016 | 13.4% | -0.061 – 0.093 | 0.334 |
|  | **Direct Effect** | -0.020 | -16.6% | -0.246 – 0.199 | 0.428 |
|  | **Total Effect** | 0.119 |  | 0.052 – 0.187 | **<0.001** |
| **Hypnogram** | Parenchymal Resistance (*Placebo*) | 0.017 | 14.4% | -0.042 – 0.077 | 0.274 |
|  | Parenchymal Resistance (*Treatment*) | 0.086 | 72.2% | -0.009 – 0.197 | **0.036** |
|  | Cerebrovascular PTT (NREM) (*Placebo*) | -0.020 | -17.2% | -0.062 – 0.008 | 0.073 |
|  | Cerebrovascular PTT (NREM) (*Treatment*) | 0.013 | 10.5% | -0.051 – 0.088 | 0.323 |
|  | N2 Sleep Duration | 0.010 | 8.0% | -0.021 – 0.046 | 0.239 |
|  | WASO Duration | 0.007 | 5.8% | -0.028 – 0.048 | 0.321 |
|  | Peripheral PTT (NREM) | 0.008 | 6.9% | -0.057 – 0.078 | 0.403 |
|  | **Direct Effect** | 0.068 | 57.2% | -0.035 – 0.174 | 0.096 |
|  | **Total Effect** | 0.119 |  | 0.051 – 0.187 | **<0.001** |

\*Effect Size, Indirect Effect, Direct Effect or Total Effect; 95% CI, 95% confidence interval. Bayes 1-Sided P values were computed from the mass of the posterior distribution that crossed zero. **Bolded P values reflect significant associations at P < 0.05**

**Supplementary Table 5:** Pharmacodynamic mediators and suppressors of glymphatic clearance engaged in the Dexmedetomidine Trial*\**.

| **Sleep Mediators in Model** | **Mediator or Suppressor** | **Effect Size** | **Fraction of Total Effect (%)** | **95% CI** | **Bayes 1-Sided P value** |
| --- | --- | --- | --- | --- | --- |
| **Slow Wave Count** | Slow Wave Count | -0.072 | -54.3% | -0.468 – 0.206 | 0.287 |
|  | Cerebrovascular PTT (NREM) | -0.082 | -61.8% | -0.357 – 0.095 | 0.142 |
|  | LFHF Ratio (NREM) | 0.019 | 14.5% | -0.106 – 0.166 | 0.351 |
|  | Resp Rate (NREM) | 0.021 | 16.1% | -0.345 – 0.409 | 0.429 |
|  | End-Tidal CO2 (NREM) | 0.039 | 29.3% | -0.225 – 0.367 | 0.321 |
|  | **Direct Effect** | 0.251 | 189.3% | -0.161 – 0.657 | 0.103 |
|  | **Total Effect** | 0.133 |  | -0.019 – 0.279 | **0.042** |
| **EEG Powerband** | EEG Slow-Delta (NREM) | -0.128 | -100.2% | -0.671 – 0.287 | 0.292 |
|  | Cerebrovasccular PTT (NREM) | -0.082 | -63.9% | -0.342 – 0.070 | 0.104 |
|  | LFHF Ratio (NREM) | 0.039 | 30.4% | -0.081 – 0.213 | 0.232 |
|  | Resp Rate (NREM) | -0.019 | -15.1% | -0.280 – 0.203 | 0.420 |
|  | End-Tidal CO2 (NREM) | 0.058 | 44.9% | -0.088 – 0.310 | 0.168 |
|  | **Direct Effect** | 0.296 | 231.1% | -0.258 – 0.838 | 0.140 |
|  | **Total Effect** | 0.128 |  | -0.020 – 0.283 | **0.044** |
| **Hypnogram** | N2 Sleep | -0.163 | -125.3% | -1.804 – 3.022 | 0.353 |
|  | Cerebrovascular PTT (NREM) | -0.091 | -70.3% | -0.836 – 0.128 | 0.145 |
|  | REM Sleep [min] | 0.129 | 99.5% | -0.427 – 0.643 | 0.190 |
|  | WASO Sleep | 0.198 | 152.3% | -1.472 – 0.824 | 0.151 |
|  | LFHF Ratio (NREM) | 0.052 | 40.1% | -0.309 – 0.594 | 0.297 |
|  | Resp Rate (NREM) | -0.051 | -39.7% | -0.426 – 0.263 | 0.322 |
|  | End-Tidal CO2 (NREM) | 0.103 | 79.7% | -0.224 – 1.194 | 0.162 |
|  | **Direct Effect** | -0.013 | -9.9% | -2.050 – 1.879 | 0.489 |
|  | **Total Effect** | 0.130 |  | -0.037 – 0.282 | 0.056 |

\*Effect Size, Indirect Effect, Direct Effect or Total Effect; 95% CI, 95% confidence interval. Bayes 1-Sided P values were computed from the mass of the posterior distribution that crossed zero. **Bolded P values reflect significant associations at P < 0.05**

## Notes

### Clinical Trial

Clinical Trial ID: NCT07432997

### Author Declarations

Western Institutional Review Board (IRB No. 20232285) gave ethical approval for this work.

### Summary of Updates

This version of the manuscript has been revised to include corrected references and additional details in the figure captions.

